# Spatial Allocation of Scarce COVID-19 Vaccines^*†‡^

**DOI:** 10.1101/2020.12.18.20248439

**Authors:** François M. Castonguay, Julie C. Blackwood, Emily Howerton, Katriona Shea, Charles Sims, James N. Sanchirico

**Affiliations:** Department of Agricultural and Resource Economics, University of California, Davis, Davis, CA 95616, USA; Department of Mathematics and Statistics, Williams College, Williamstown, MA 01267, USA; Department of Biology and Center for Infectious Disease Dynamics, Pennsylvania State University, University Park, PA 16802, USA; Howard H. Baker Jr. Center for Public Policy and Department of Economics, University of Tennessee, Knoxville, Knoxville, TN 37996, USA; Department of Environmental Science and Policy, University of California, Davis, Davis, CA 95616, USA and Resources for the Future, Washington DC 20036, USA

**Keywords:** economic epidemiology, applied numerical methods, efficient disease intervention, allocation of scarce resources

## Abstract

The COVID-19 Vaccines Global Access (COVAX) is an initiative led by the World Health Organization (WHO) and other partners that aims for an equitable access of COVID-19 vaccines. Despite a potential heterogeneous disease burden across space, countries receiving allotments of vaccines via COVAX may want to follow WHO’s allocation rule and distribute vaccines to their jurisdictions based on the jurisdictions’ relative population size. Utilizing economic–epidemiological modeling, we benchmark the performance of this *ad hoc* allocation rule by comparing it to the rule that minimizes the economic damages and expenditures over time, including a penalty cost representing the social costs of deviating from the *ad hoc* allocation. Under different levels of vaccine scarcity and different demographic characteristics, we consider scenarios where length of immunity and compliance to travel restrictions vary, and consider the robustness of the rules when assumptions regarding these factors are incorrect. The benefits from deviating are especially high when immunity is permanent, when there is compliance to travel restrictions, when the supply of vaccine is low, and when there is heterogeneity in demographic characteristics. Interestingly, a lack of compliance to travel restrictions pushes the optimal allocations of vaccine towards the *ad hoc* and improves the relative robustness of the *ad hoc* rule, as the mixing of the populations reduces the spatial heterogeneity in disease burden.

**JEL Classification:** C61, H12, H84, I18, Q54

## 1 Introduction

Now that several vaccines against coronavirus disease 2019 (COVID-19) have obtained the World Health Organization’s (WHO) emergency use listing, policymakers around the globe are deciding how to allocate the limited supplies within their boundaries. While the vaccination campaign of developed countries like the U.S. and the U.K. is well under way, developing countries in Asia and Africa face major vaccine supply issues. In anticipation of this problem, WHO and other partners created “COVID-19 Vaccines Global Access” (COVAX)—an initiative that aims for an equitable access of COVID-19 vaccines. The scientific literature has addressed the question of vaccine allocation, but most of it has focused on demographic considerations within one jurisdiction [1; 2; 3] or on a global scale [4; 5; 6; 7]. This prior work has made important contributions to the debate. A missing piece in the allocation question, however, is how to divide up limited quantities across jurisdictions (e.g. state/provinces, counties/regions) that might have different demographic and epidemiological characteristics. In the U.S., the National Academies of Sciences, Engineering, and Medicine (NASEM) recommends that vaccines are allocated to jurisdictions based on their relative population size “in the interest of speed and workability” [8] and WHO applies a similar principal with its COVAX program [9]. Countries whose vaccination campaign depends on COVAX provision could also apply a similar principle within their boundaries.

In this paper, we derive the optimal allocation and investigate its robustness to incorrect behavioral (i.e. compliance to nonpharmaceutical interventions) and epidemiological (i.e. the duration of immunity) assumptions. We also explore the economic and epidemiological trade-offs associated with a fixed *ad hoc* allocation rule similar to NASEM’s [8] and WHO’s [9] principle by comparing it to the optimal rule conditional on the level of scarcity of the vaccine.

Furthermore, we investigate how various vaccine effectiveness—mimicking the wide range of effectiveness of the vaccines available for distribution (for the lower and upper end of the spectrum, see [10; 11] respectively) and the reduced effectiveness of vaccines when faced with emerging variants of concern [12]—affect the optimal allocation. The optimal rule we consider minimizes the economic costs from health-related damages, vaccine expenditures, and a workability cost imposed on the planner for deviating from the *ad hoc* rule.

In a world where two jurisdictions are identical in terms of population, the *ad hoc* rule would divide the limited supply equally between the jurisdictions. However, it is much more likely that two jurisdictions, even if equally sized, have heterogeneous levels of infections (e.g. in terms of cases) at the time a country’s central government receives an allotment of vaccines. Based on prior literature on spatial-dynamics of disease management, heterogeneity in infection levels may lead to significant deviations between the optimal spatial allocation and the *ad hoc* rule potentially leading to greater economic costs and worse public health outcomes (see [13] for example).

Mechanisms leading to heterogeneous infection include the timing of the outbreak, demographic characteristics of the population (e.g. age structure [14] and essential worker status [15]), and the implementation of and compliance with preventative nonpharmaceutical interventions; see [16; 17] for more details on how SARS-CoV-2 (i.e. the virus that causes COVID-19) prevalence varies across space in the U.S. and see [18] for India, where the cumulative number of cases is highly heterogeneous across states. While compliance to preventive measures may seem independent from vaccine allocation, it affects the initial conditions (i.e. the conditions before the vaccine is allotted to the country) and the conditions under which the limited supplies will be allocated within the country. For example, compliance to shelter-in-place and travel restrictions results in little to no movement of the virus from one jurisdiction to another. When regions are non-interacting, Brandeau et al. [19] show for a general susceptible–infected–susceptible (SIS) model that the optimal allocation of resources depends on numerous intrinsic factors, including the size of the populations of each jurisdiction and the level of infection at the time of the vaccine allotment. When regions are interacting, Rowthorn et al. [20] show when there is no immunity (i.e. in an SIS model) that treatment should be preferentially directed towards the region that has the lower level of infection. While these results indicate that a fixed *ad hoc* rule is less cost-effective in an SIS model, whether compliance to travel restrictions makes the *ad hoc* rule relatively more cost-effective in the case of COVID-19 is an open question.

Our findings illustrate that the vaccines should be optimally allocated over time depending on: (i) if the jurisdiction has a lower or higher disease burden at the time of the vaccine allotment, (ii) if immunity is permanent (see Zhou et al. [21]) or temporary (Gersovitz and Hammer [22] already pointed out that the optimal allocation is conditional on the duration of immunity), (iii) whether there is compliance to travel restrictions or not, (iv) the amount of vaccine available, and (v) the average demographic characteristics of the population (i.e. age structure and essential worker status). We proxy variability in demographics by assuming that the population of one jurisdiction has a higher case-fatality ratio (e.g. an older population [14]) or a higher contact rate (e.g. a population with more essential workers [15]) than the other. We find that the benefits of deviating from the *ad hoc* rule—which prioritizes equity of distribution—are especially high when immunity is permanent, when there is compliance to travel restrictions, when the vaccine supply is low, and when there is heterogeneity in demographic characteristics. Allocating a vaccine based on an *ad hoc* allocation rule generally leads to an over-utilization in jurisdictions where disease prevalence is higher, an under-utilization in jurisdictions where disease prevalence is lower, and overall a higher number of cumulative cases. Whether these inefficiencies outweigh the “speed and workability” [8] inherent in *ad hoc* rules is an important question for policymakers. Our research can aid in that discussion by illuminating the trade-offs involved in such complex epidemiological, economic, and social decisions by providing optimal benchmarks from which to compare *ad hoc* rules.

While the optimal allocation is conditional on a number of factors mentioned above, the science remains unresolved on the duration of immunity to SARS-CoV-2, and it is difficult to anticipate and subsequently estimate the extent to which populations in different jurisdictions comply with the travel restrictions. On the other hand, the *ad hoc* allocations have the advantage of being based on easily observable factors (e.g. a jurisdiction’s population size). To gain insights into the robustness of optimal and *ad hoc* policies in the presence of such uncertainties, we investigate the economic and public health consequences that could occur if we design an optimal policy or evaluate the performance of *ad hoc* rules under a set of assumptions on immunity and compliance that turn out to be incorrect. We make a number of contributions to the literature. First, we develop an economic–epidemiological model and solve for the optimal allocation of vaccines over time to minimize the economic costs from health-related damages, vaccine expenditures, and a workability cost imposed on the planner for deviating from the *ad hoc* rule. Prior literature considering the trade-offs involved with *ad hoc* rules does not consider that deviating from them entails potential workability costs (see, for example, [23]). Second, we consider how vaccine allocations are influenced by compliance with preventative nonpharmaceutical interventions (i.e. travel restrictions). Third, we demonstrate how vaccine allocations are dependent on various demographics (i.e. age structure and essential worker status). Fourth, we show that, in general, optimal rules are robust to incorrect assumptions about the duration of immunity but differences in public health outcomes (cumulative cases) appear when compliance to a travel restrictions is assumed when in fact there is not compliance; it is, however, much preferable from a public health outcome perspective to comply with travel restrictions.

The paper is divided as follow. In Section 2, we detail the different types of interventions, we present the components of the economic-epidemiological model, and detail the technique used to analyse the allocation question. Section 3 presents the results while Section 4 concludes the paper.

## 2 Material and Methods

We develop an economic–epidemiological model to describe the dynamics of SARS-CoV-2. The model captures a situation where a central planning agency (e.g. the central government) must decide when and how much of the scarce vaccines to allocate to two jurisdictions where disease burden is heterogeneous at the moment it receives an allotment of vaccine. We assume that the objective of the central planner is to minimize costs across both jurisdictions, including damages associated with the morbidity and deaths of infected individuals, the expenditures related to the pharmaceutical intervention, and a penalty cost mimicking the increased workability costs incurred for any deviation from the *ad hoc* allocation. The dynamics of SARS-CoV-2 are modeled using an SEIR epidemiological model, which tracks the change over time of the susceptible (S), exposed (E), infected (I), and recovered (R) populations for two separate jurisdictions (see Appendix A for more details on the calibration of the model). We note that while we generally talk about these jurisdictions as being two different states, they can very well represent any two sub-national jurisdictions like provinces or territories, and even counties or regions within one sub-national jurisdiction.

### 2.1 Modelling Different Types of Intervention

There are two different types of interventions we consider: travel restrictions and vaccines. We assume that travel restrictions affect both jurisdictions simultaneously (e.g. by an order from the central government), and that the populations either comply perfectly or imperfectly to the travel restrictions (for examples of optimal lockdown policies see, e.g., [24; 25]). When compliance is perfect, individuals in different jurisdictions do not interact with each other and thus susceptible individuals can only get infected by being in contact with some infected individual in their own jurisdiction. When compliance is imperfect, susceptible individuals from one jurisdiction can also travel to the other jurisdiction where they can be in contact with infected individuals, or infected individuals from one jurisdiction can travel to the other jurisdiction and infect susceptible individuals there; this discrete shift in the number of contacts effectively increases the transmissibility of the virus (see Appendix A more details).

We assume that the analysis starts when the central planning agency has received an allotment of vaccines and will keep receiving a continuous allotment of vaccines. For simplicity, the amount of available vaccine is assumed to be exogenous to the model and fixed over time, which is likely given the short time frames we consider in the paper. However, we consider different levels of vaccine allotments, or capacity, to investigate how different levels of vaccine scarcity may affect their optimal allocation. In our model, vaccines reduce the pool of susceptible individuals by providing them with immunity from the virus, as early evidence suggests that vaccines could be transmission blocking in addition to preventing severe disease [26].

### 2.2 Model of Disease Transmission

We use a frequency-dependent [27] susceptible–exposed–infected–recovered (SEIR) model that describes the dynamics of COVID-19 in two separate jurisdictions *i* = 1, 2 (e.g. states/provinces or counties/regions); each jurisdiction contains a population of *N*_*i*_ individuals that is either susceptible, exposed, infected, or recovered (see Figure 1). We also consider scenarios where immunity is temporary (i.e. lasts 6 months, for more details see [28]), thus also using an SEIR–Susceptible (SEIRS) model (for COVID-19 applications see, e.g., [29; 30; 31; 32; 33; 34; 35; 36; 37; 38; 39]). In such scenarios, the *R*_*i*_ recovered individuals are immune for a mean period of 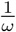 months.

**Figure 1:**
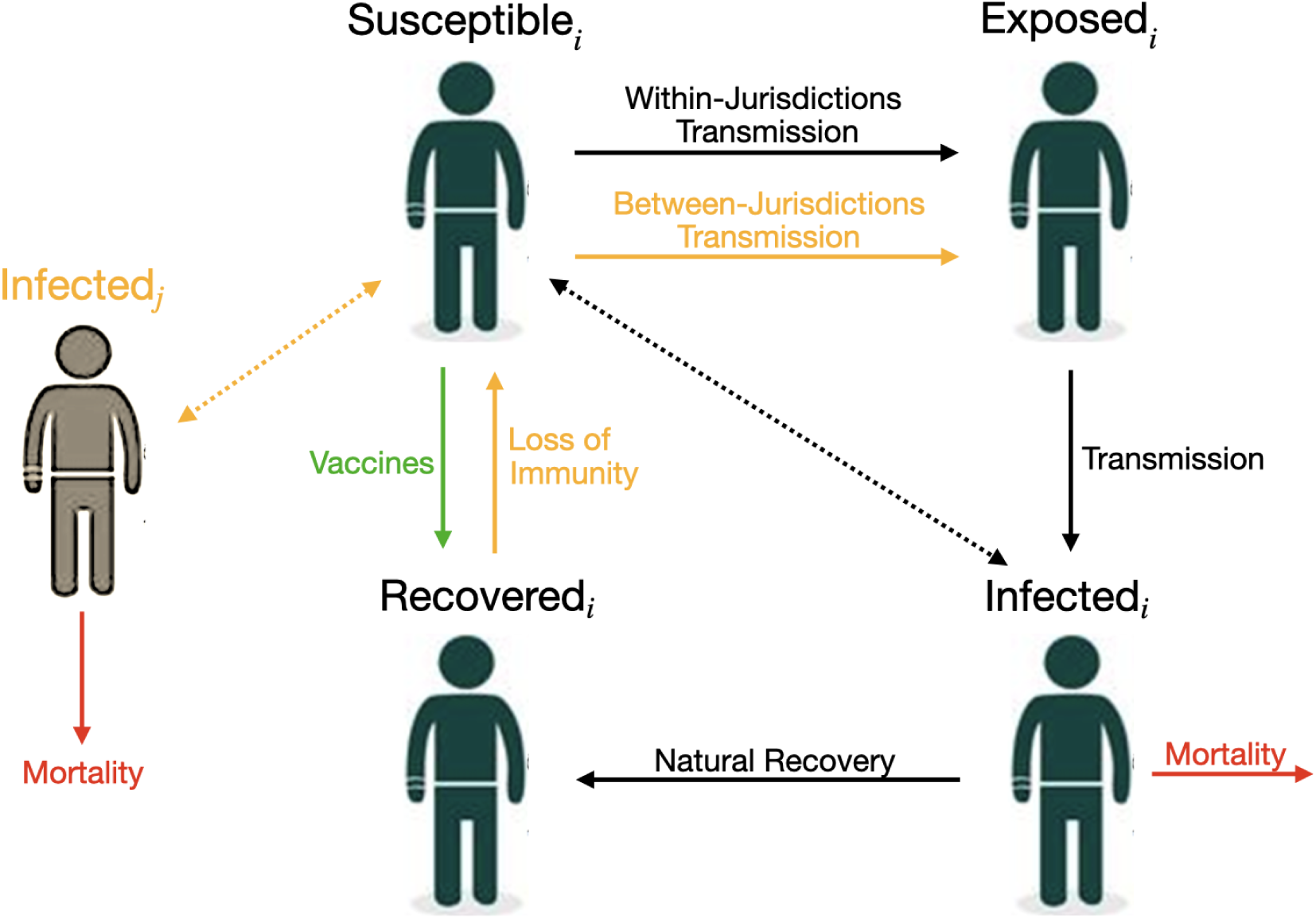
This schematic shows the model interventions and disease transmission pathways for our model of COVID-19. The full lines represent the transition between, or out of, compartments while the dotted lines represent contact between susceptible and infected individuals. Black lines represent situations that do not vary, while yellow lines represent key factors that we vary in our model to see how they impact our results. The green line represents the vaccines and the red line represents mortality.

In each jurisdiction *i*, the *S*_*i*_ susceptible individuals are in contact with the *I*_*i*_ infected individuals of their own jurisdiction at a rate of *β*_*ii*_ and are in contact with the *I*_*j*_ infected individuals of the other jurisdiction at a rate of *β*_*ij*_. We assume *β*_*ij*_ = 0 (i.e. no mixing between jurisdictions) when there is perfect compliance to travel restrictions, and *β*_*ij*_ *>* 0 if not. To highlight the role of travel restriction compliance and initial disease burden, we initially assume that the contact rate is identical across jurisdictions, meaning that *β*_11_ = *β*_22_ = *β*_*ii*_ and *β*_12_ = *β*_21_ = *β*_*ij*_ (in Section 3.2 we relax this assumption and investigate the optimal allocation when there is heterogeneity in the contact rate). We assume there is no permanent migration of individuals from one jurisdiction to another (see for instance [40] and see [41] for an example applied to COVID-19) in the sense that individuals who do not comply with travel restrictions do not permanently move to the other state, but instead travel to it temporarily. An implication is that we are assuming that the two jurisdictions are close enough for such travel and mixing to be economically feasible.

We model the control variables for vaccines as non-proportional controls, i.e. available in a constant amount each month [3; 20; 42; 43]. The change in susceptible individuals is

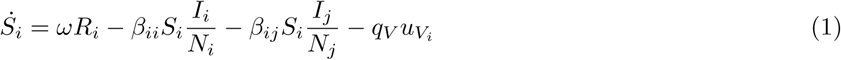

where *u*_*V*_*i* represents the number of individuals being treated via vaccine in a given time period (i.e. a month) in Jurisdiction *i*, and *q*_*V*_ represents the effectiveness of the vaccine (note that we look at the low end of vaccine effectiveness to be conservative; see Section 3.4 for more details). We note that our model does not distinguish between individuals whose vaccine has failed and those who have not been vaccinated at all. As such, individuals with vaccine failure can be re-vaccinated in subsequent months.

After being infected, susceptible individuals transition into the exposed class *E*_*i*_ where the disease remains latent for a mean period of time of 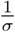, before the onset of infectiousness. The change in the number of exposed individuals is

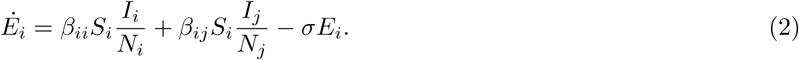

Exposed individuals eventually become infectious for a mean period of time of 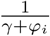 and in turn can infect susceptible individuals. Infected individuals either recover naturally from the disease at a rate of *γ* or die from complications related to infection at a disease induced mortality rate of *φ*_*i*_. In our base case we assume identical disease induced mortality rates across jurisdictions, i.e. *φ*_1_ = *φ*_2_ = *φ* but investigate the optimal allocation when *φ*_1_ = *φ*_2_ in Section 3.2. The growth of the infected individuals is

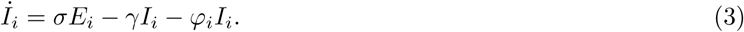

The recovered population *R*_*i*_ includes individuals that recover naturally from the disease at a rate of *γ* and the individuals that are successfully vaccinated every month 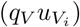; if immunity is temporary (*ω >* 0), a fraction of the recovered will leave this compartment. Our model does not distinguish between vaccine-acquired immunity and naturally-acquired immunity. The number of recovered individuals in Jurisdiction *i* thus changes according to

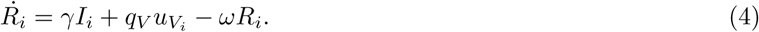

At any instant in time, we have that *N*_*i*_ = *S*_*i*_ + *E*_*i*_ + *I*_*i*_ + *R*_*i*_, which in turn implies that the growth of the population over time is

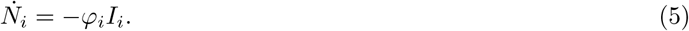

In keeping with much of the previous economic epidemiology literature [22] as well as recent applications to COVID-19 (see for example [44]), we have omitted natural births and non-COVID-related deaths due to the short time frame of our model (4 months) and assume reductions in international travel [45] effectively lead to a closed population (i.e. there is no exogenous importation of infected individuals). See Appendix A for more details about the parameterization of the epidemiological model.

### 2.3 Modelling *Ad Hoc* Allocations

We model an *ad hoc* allocation rule that favors “speed and workability” [8]. We follow the NASEM [8] and WHO [9] principle and impose that the allocation is based on relative population sizes. Specifically, the rule for Jurisdiction *i* is that

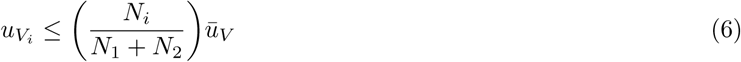

where *ū*_*V*_ is the limited amount of vaccine allotted to the central government. When the population sizes are the same, the *ad hoc* rule will divide equally the limited doses to the two jurisdictions.

In the *ad hoc* scenarios, we model the allocation rule as an inequality because towards the end of the horizon after periods of vaccinations, the level of susceptible in the population may be such that the limited supply of vaccines is not an issue. Other *ad hoc* rules are possible, such as, allocate all to the largest or smallest population [23], but we concentrate on the one currently being advocated for by NASEM [8] and WHO [9].

### 2.4 Model of Economic Costs

The model of economic costs include damages related to morbidity and deaths, costs spent on the vaccines, and the workability cost described above that is incurred for any deviation from the *ad hoc* allocation rule. Damages represent consequences related to a temporary disability associated with severe or critical symptoms, and loss of life in the worst cases. The damages are assumed to be linear and additively separable across jurisdictions, meaning that they are identical across individuals and across jurisdictions. The marginal value of damages (i.e. the damages associated with the death of one individual) is assumed to be constant over time and given by the value of a statistical life (VSL) that the U.S. Environmental Protection Agency [46] uses (see Appendix A for more details on the parameterization). Damages incurred from a temporary disability associated with severe or critical symptoms can be compared to deaths via some disability weight *w*; given we found no published disability values associated with COVID-19, we follow the literature (see for instance [47]), and use the disability value associated with lower respiratory tract infections. The damage function for Jurisdiction *i* is

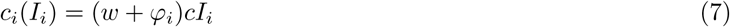

where *c* is the damage parameter associated infectious individuals (i.e. the VSL).

We model a scenario where the central planner is focused on the allocation of vaccines where the costs for its development have already been incurred. This implies that vaccine development costs have already been utilized (in technical terms we say that the costs are sunk) and therefore do not affect the decision of the central planning agency. We model the vaccination cost as linear, where the cost parameter represents the cost of vaccinating one individual. The vaccine cost function is denoted 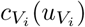, with *i* = 1, 2. We assume that the vaccination cost is additively separable across jurisdictions such that we denote the cost of vaccinating 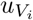 individuals as

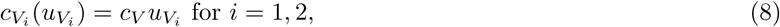

where *c*_*V*_ represents the cost of treating one individual via vaccine. Note that calibration of the cost parameter is based on current vaccine prices (see Appendix A for more details about the parameterization of the economic model), but it could also represent the cost the central planner needs to incur to pay to administer the vaccine.

We assume that the central planning agency incurs a workability cost representing the social (transaction) costs of deviating from the *ad hoc* allocation rule (for another application of this concept, see [48]). The workability cost function is:

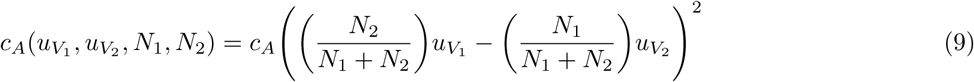

where *c*_*A*_ is the parameter associated with the workability cost. When the gains from deviating from the *ad hoc* allocations (i.e. a reduction in damages in one jurisdiction) outweigh the costs (i.e. an increase in damages in the other jurisdiction and the increased workability costs incurred), the central planning agency will prioritize this allocation as it will lead to lower total costs. By imposing the *ad hoc* rule *ex ante*, the decision-maker is essentially assuming that this workability cost is infinite. Everything else being equal, we expect that the presence of the workability cost will push the optimal allocation towards the *ad hoc* rules (see Figure A18 for a sensitivity analysis of our results to the workability cost parameter). Therefore, when we do find deviations, we need to consider that these include this workability cost and if workability costs smaller, then the deviations and trade-offs would be greater.

### 2.5 Planner’s Objective

In optimal control theory, the best, or optimal, path of the control variables (here the allocation of the limited supply of vaccines) is conditional on the objective of the central planning agency. We assume that the objective is to minimize the economic damages and the costs of the pharmaceutical intervention across jurisdictions over time, rather than a solely epidemiological objective (see for instance [20]). The objective function is the net present value of damages, expenditures related to vaccination, and the workability cost over an exogenously determined planning horizon (4 months). Specifically, the planner’s objective is:

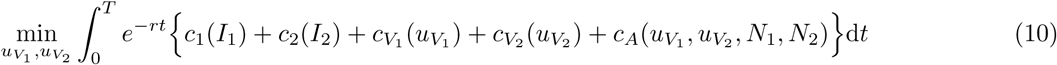

where *r* is monthly discount rate. The planner solves equation (10) over a fixed time interval, *T*, subject to equations (1), (2), (3), (4), (5), along with constraints on availability of vaccines 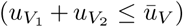, non-negativity conditions, physical constraints on vaccines, initial disease burdens in each jurisdictions, and free endpoints (see discussion on terminal conditions in the next section). In the *ad hoc* scenarios, we also impose equation (6).

### 2.6 Initial and Terminal Conditions

The disease burden in each jurisdiction at the beginning of the time horizon (i.e. in *t* = 0 when the allotment of vaccine is received for the first time) is calibrated using the epidemiological model (equations (1), (2), (3), (4), and (5)). At the beginning of the outbreak, we assume that, in each jurisdiction, there is one exposed individual in an otherwise entirely susceptible population of 10 million individuals, and that populations of the different jurisdictions comply with the travel restrictions. The only difference between the two jurisdictions is that the outbreak started one week earlier in State 2. We simulate the outbreak for approximately nine months to yield the initial conditions; see Appendix B for more details. In Section 3.2 when we consider heterogeneity in demographic characteristics (varying case-fatality ratio and contact rate), we modify the initial conditions accordingly assuming an identical timing in the outbreak of the disease.

We impose no conditions on the number of susceptible, exposed, infected, and recovered individuals at the end of the planning horizon; in technical terms, we say that the state variables are free (see Appendix B for more details). Under our free endpoint conditions, there is a transversality condition (i.e. a necessary condition for the vaccine allocation to be optimal) for each state variable that requires the product of the state variable (*S*_*i*_, *E*_*i*_, *I*_*i*_, *R*_*i*_ or *N*_*i*_) and its corresponding costate variable (i.e. the shadow value, or cost, associated with the state variable) is equal to zero. Hence, at the end of the time horizon, either the state variable equals zero, the shadow value associated with the state variable equals zero, or both. In any case, allowing state variables to be free guarantees that the terminal levels of the state variables are optimally determined. Another possible assumption could be that over a fixed interval we find the optimal policy such that at the end of the horizon there is a given percent reduction in infected or susceptible individuals. Our approach nests this more restricted scenario.

## 3 Results

To examine the optimal allocations of vaccine over time, we numerically solve the optimal control problem across three different scenarios: no controls, optimal vaccine allocation, and *ad hoc* vaccine allocation. We investigate how to allocate vaccines by mapping out the different allocation rules for different immunity–travel restrictions–capacity scenarios. Any deviation from the *ad hoc* allocation rule is optimal despite incurring the workability cost. As the workability cost parameter *c*_*A*_ goes to zero, the problem becomes linear in the controls where the optimal allocations in linear problems follow singular solutions. We use pseudospectral collocation to solve for the optimal dynamics of vaccine and infection over time, which converts the continuous time optimal control problem into a constrained non-linear programming problem solving for the coefficients of the approximating polynomials at the collocation nodes (see [49; 50] for other applications, and see Appendix B for more details on this technique).

We present the results for our preferred specification of the parameters (i.e. following what was estimated in the literature; see details in Appendix A) and for the case where immunity is permanent and the case where immunity is temporary. We detail the optimal deviation based on whether the populations of the different jurisdictions are compliant to travel restrictions or not, and for different levels of capacity constraints. The total available quantity of vaccine in a given time period (i.e. a month; *ū*_*V*_) is based on a certain percentage (5%, 10%, or 15%) of the total population size. We focus our analysis on the period of time when the scarcity of the vaccine constraint is binding, as once the constraint relaxes the allocation question becomes moot.

### 3.1 Base Case: Homogeneous Demographic Characteristics

Compliance to travel restrictions impacts the optimal allocation of vaccines, regardless of whether immunity is temporary or permanent and regardless of the amount of vaccine available. Noncompliance to travel restrictions reduces both the oscillation (i.e. back-and-forth movement of resources between jurisdictions) of the optimal allocation and the amplitude of the deviations from the *ad hoc* rule (see Figure 2 for when immunity is permanent, and see Figure A1 for when immunity is temporary). Because noncompliance to travel restrictions decreases the structural heterogeneity in the system, the optimal allocation of vaccine converges towards the *ad hoc* allocation when populations mix with each other. This result clearly demonstrates how the performance of the allocation rule is dependent on how citizens in the jurisdictions comply with travel restrictions.

**Figure 2:**
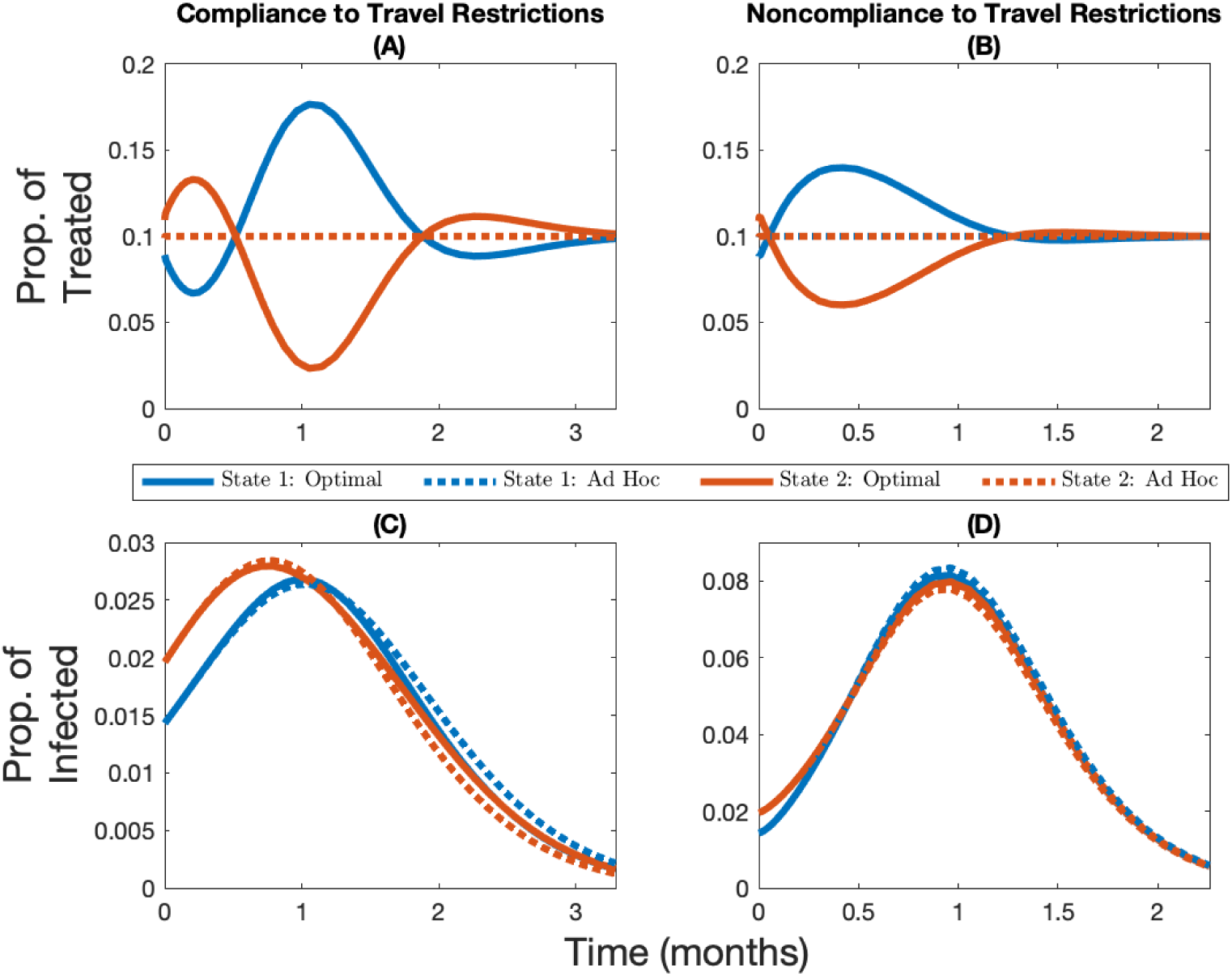
Vaccine allocation with and without compliance to travel restrictions. Change over time in the optimal and *ad hoc* allocations (panels A and B) and the corresponding infection levels (panels C and D) for State 1 (in blue, the initially lowest-burdened state) and State 2 (in red, the initially highest-burdened state) depending on whether there is compliance to travel restrictions (panels A and C) or not (panels B and D) for the case where the vaccine capacity constraint is 10% and immunity is permanent. Note the changing *y*-axis in panels C and D in order to better highlight the infection levels.

Noncompliance to travel restrictions leads to the initially less infected state being favored by the optimal allocation for low levels of vaccine capacity (e.g. 5% capacity; see Figure 3 Panel A for when immunity is permanent and Figure A4 Panel A for when immunity is temporary). On the other hand, the more infected state will be prioritized at the beginning of the time horizon for a very short period of time when vaccine capacity is larger (e.g. 10% or 15% capacity; see Figure 3 panels B and C for when immunity is permanent and Figure A4 panels B and C for when immunity is temporary). More generally, regardless of whether or not populations are compliant with travel restrictions, and regardless of whether immunity is temporary of permanent, a higher vaccine capacity implies that relatively more of the supply should be given to the more infected state at the beginning of the time horizon (see figures 3 and A2 for the case where immunity is permanent; see figures A3 and A4 for the case where immunity is temporary).

**Figure 3:**
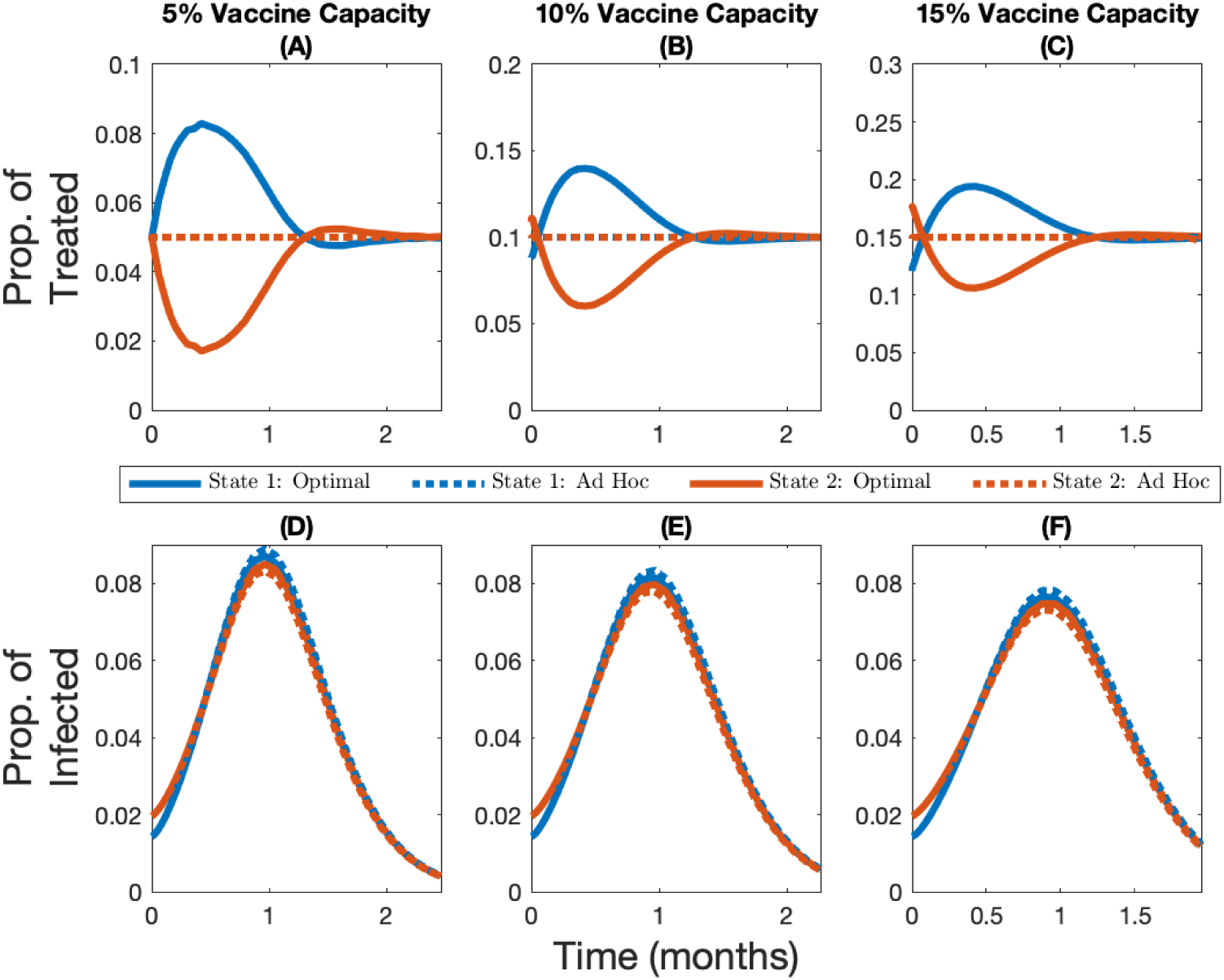
Vaccine allocations under different levels of scarcity without compliance to travel restrictions. Change over time in the optimal and *ad hoc* allocations (panels A, B, and C) and the corresponding infection levels (panels D, E, and F) for State 1 (in blue, the initially lowest-burdened state) and State 2 (in red, the initially highest-burdened state) depending on whether capacity is 5% (panels A and D), 10% (panels B and E), or 15% (panels C and F), for the case where immunity is permanent and there is no compliance to travel restrictions.

Interestingly, temporary immunity has a different effect on the optimal vaccine allocation depending on whether or not populations are compliant to travel restrictions. When populations comply with travel restrictions, temporary immunity increases the oscillation of the optimal allocation because benefits from vaccination are only temporary, and since the population gradually loses its immunity, it forces more back-and-forth movement of resources between jurisdictions (see Figure A5). When populations do not comply with travel restrictions, temporary immunity reduces the amplitude of the deviations from the *ad hoc* rule because it further dampens the structural heterogeneity in the system, since the infection and recovery level of both jurisdictions will eventually reach the same positive steady-state level (recall the only heterogeneity in the system is the initial disease burden in the base case).

While the optimal allocation of vaccine is unequal from a resource allocation perspective, it equalizes the current infection levels across jurisdictions (Figure 2 Panel C), that is, the optimal allocation seems to be prioritizing equity of outcomes as opposed to the *ad hoc* allocation, which prioritizes equity of distribution. As the vaccine capacity increases, however, the *ad hoc* allocation rule performs better and in turn the amplitude of the optimal deviation decreases (see Figure A2 panels A, B and C, or Figure A3 panels A, B, and C). These optimal cost-minimizing deviations that lead to equal current infection levels across jurisdictions towards the end of the time horizon imply that the optimal cumulative number of cases is more unequal than in the *ad hoc* allocation (Figure 4). Hence, the optimal allocation makes the current infection level more equal, while the *ad hoc* allocation makes cumulative infection more equal. In fact, in all scenarios considered, the optimal allocation will lead to lower cumulative damages in the less infected jurisdiction but higher cumulative damages in the most infected jurisdiction (see Figure 4 with vaccine capacity of 10%, and see figures A7 and A8 with vaccine capacity of 5% and 15%, respectively).

**Figure 4:**
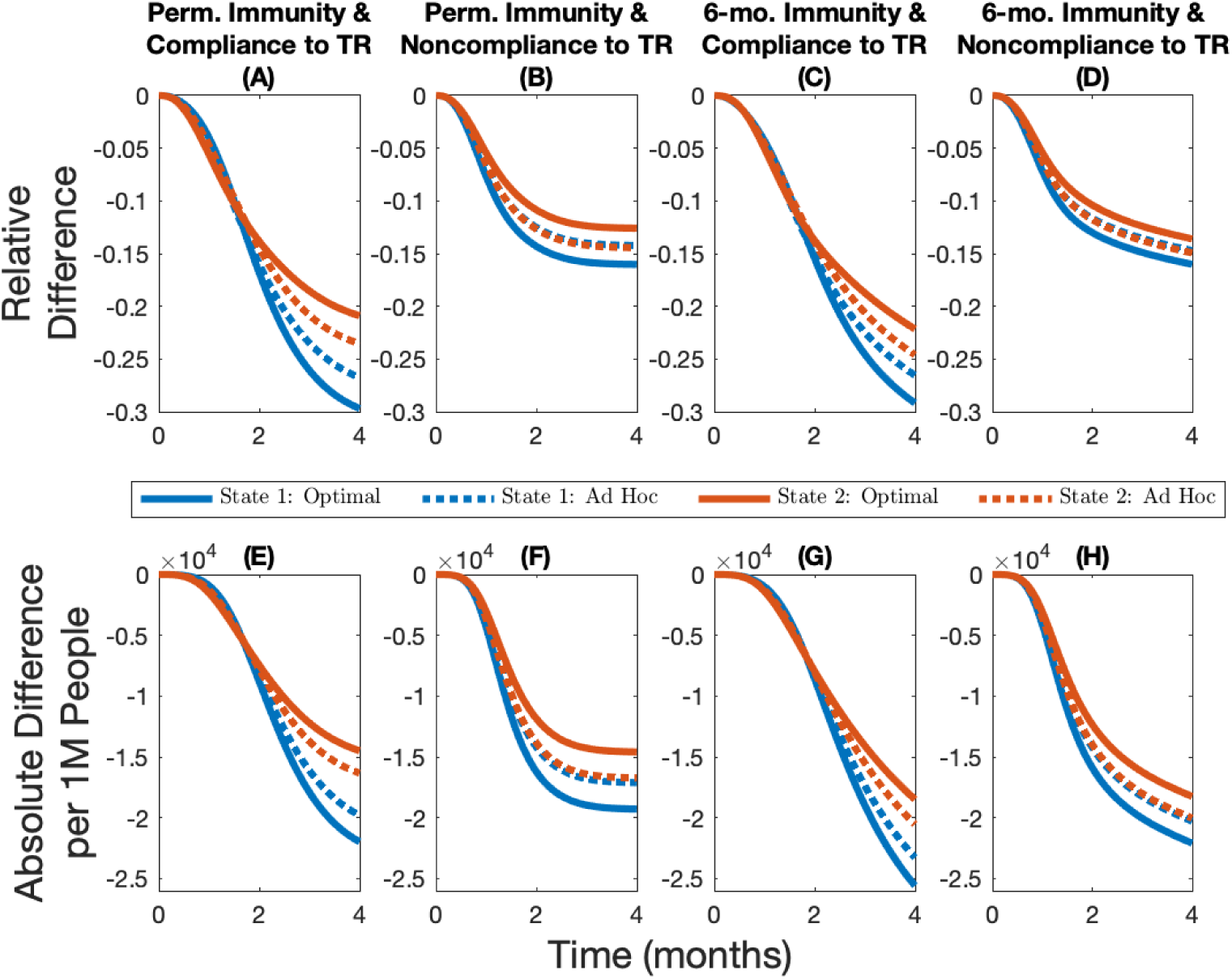
Epidemiological outcomes under different scenarios with vaccines. Cumulative relative difference (panels A, B, C, and D) and cumulative absolute difference per 1M people (panels E, F, G, and H) between the number of infections in different allocations rules and the no-vaccine case for different immunity–travel restrictions (TR) scenarios and for when vaccine capacity is 10%.

### 3.2 Heterogeneous Demographic Characteristics

In our main analyses, we introduced heterogeneity in infection across jurisdictions by assuming a different timing in the outbreak of the virus. While this leads to heterogeneity in the disease burden, there are other mechanisms that could lead to similar differences in the disease burden at the time the allotment of vaccine is received. For instance, in jurisdictions that have an older population on average, we expect SARS-Cov-2 to have a higher case-fatality ratio [14] which would effectively decrease the transmissibility of the virus (see Appendix A more details). If we assume that one jurisdiction has a higher case-fatality ratio and start the initial outbreak at the same time, then we find the population with the highest case-fatality ratio also has the largest population of susceptible individuals at the time the vaccine is administered. The heterogeneity in initial disease burden stems from the lower transmissibility in the jurisdiction with the higher case fatality ratio. The heterogeneity in case fatality ratio therefore not only leads to heterogeneity in the initial disease burden but it also implies that the benefits of vaccination are no longer homogeneous across jurisdictions. These differences lead the optimal allocation to favor even more the least burdened jurisdiction, which is also the most vulnerable (aka more older individuals) of the two populations. Overall, introducing heterogeneity in case fatality ratio strengthens our main set of results (see Figure A9 for when immunity is permanent and see Figure A10 for when immunity lasts 6 months).

Another source of heterogeneity in infection could stem from one jurisdiction having more essential workers than the other (for more details on how the risk of infection is occupation-dependent, see [15]). In our model, we can capture this by considering spatial heterogeneity in the contact rate, where a higher contact rate proxies for more essential workers. This in turn leads to a higher initial disease burden in the jurisdiction with higher contact rate. We find that priority is given to the state with a higher contact rate (aka more essential workers) in almost all cases. As the state with a higher contact rate gets vaccinated, we eventually shift priority to the state with the lowest contact rate because either the number of cases starts decreasing in the jurisdiction that has the higher contact rate, or the low-contact state eventually reaches a point where its infection level becomes higher than the high-contact state (see Figure A11 for when immunity permanent and see Figure A12 for when immunity lasts 6 months). In the case when immunity lasts 6 months and there is compliance to travel restrictions (Figure A12, Panel A), we find that priority is given to the low-contact jurisdiction, which contradicts many notions of fairness associated with vaccine allocation. With the gains of vaccination temporary and no movement of people, it turns out that the greatest return per vaccination is in the place where we can best avoid future cases (low contact rate jurisdiction). This prioritization is only fleeting however and there is more back-and-forth movement of resources between jurisdictions in this case, even though the workability cost is being incurred each time.

### 3.3 Robustness of Spatial Allocations

There is significant uncertainty associated with the duration of immunity (i.e. if it is permanent or temporary) and to what extent populations comply with travel restrictions. One argument for the *ad hoc* allocation is that uncertainty in these parameters makes the optimal allocation impossible to achieve. This uncertainty is not yet resolved and public health officials have to choose vaccine allocations based on potentially incorrect assumptions. We compare the robustness of the optimal spatial allocation to the *ad hoc* allocation. By definition, the optimal allocation minimizes the net present value of the health-related damages and total expenditures (including vaccine expenditures and the workability cost incurred because of the deviations from the *ad hoc* allocation), and thus cannot do worse on this dimension than the *ad hoc* allocation. We measure robustness by first inserting the optimal solution under one set of assumptions into the disease dynamics under another set and compute the changes in total expenditures (i.e. the pharmaceutical intervention and the workability cost) and public health outcomes (cumulative cases) over time. We then calculate the distance of these changes in percentage terms to the optimal solution derived under the “correct” assumptions (represented by the point (0, 0) in Figure 5). For example, suppose immunity is permanent and there is perfect compliance to a travel restriction. We derive the optimal policy under these assumptions and use it to measure the robustness of the optimal policies that are derived under assumptions that immunity is temporary and/or there is noncompliance. The *ad hoc* policies being based on observable factors are then compared to the incorrectly applied optimal policies. We illustrate the case for 10% scarcity and include other scarcity cases in Appendix C.

**Figure 5:**
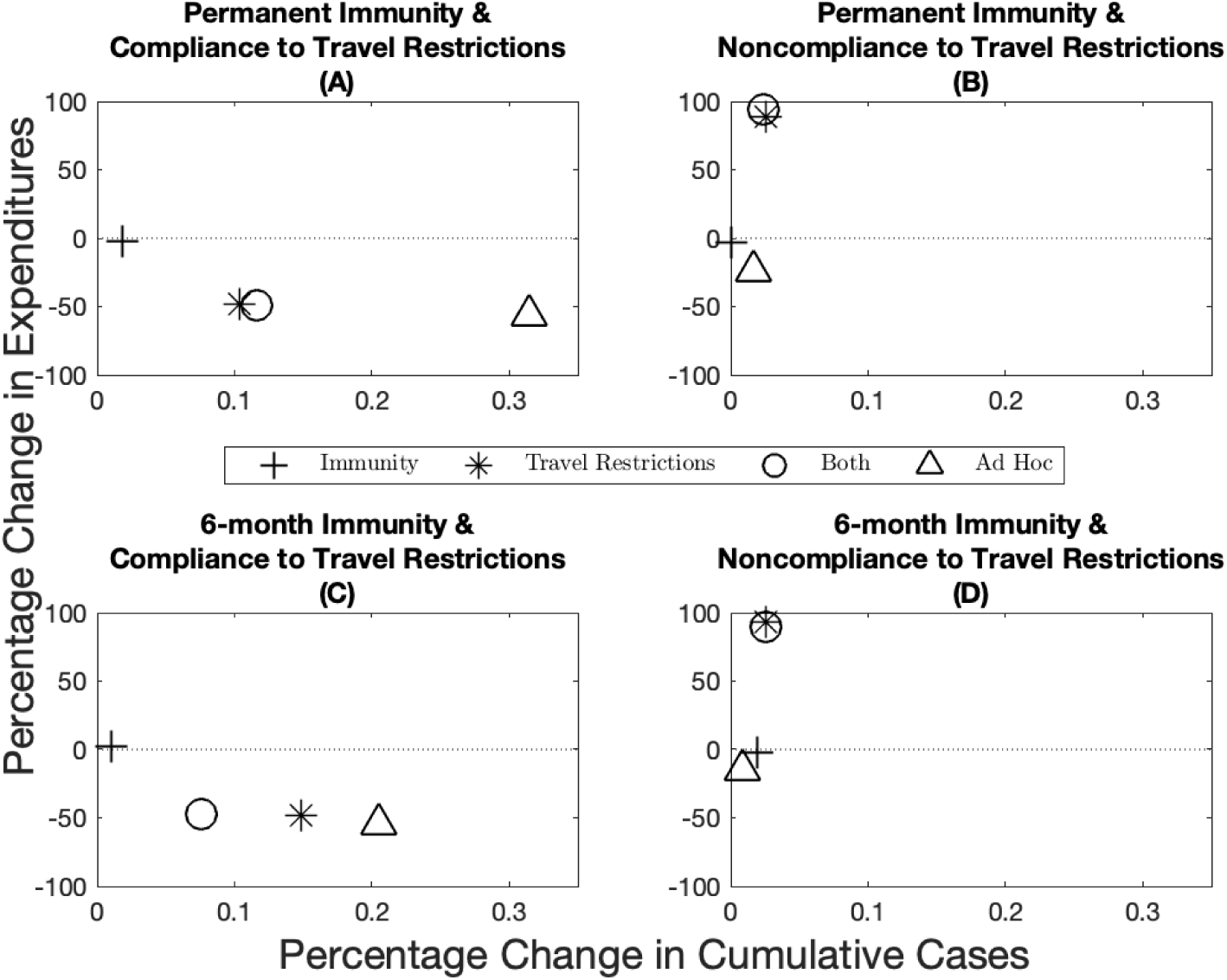
Robustness of epidemiological and economic outcomes under different scenarios with vaccines. Percentage change in expenditures (*y*-axis) and percentage change in cumulative cases (*x*-axis) from the optimal allocation for different immunity–travel restrictions scenarios and for when vaccine capacity is 10%. The *x*-axis represent small percentage changes but when scaled up to population level effects translate into significant differences in public health outcomes.

When demographic characteristics are homogeneous across jurisdictions, we find overall that immunity length has a lesser impact on both economic and epidemiological outcomes than compliance to travel restrictions (compare the distance from the origin between the plusses and the stars in Figure 5). There are more nuanced trade-offs, however (e.g. compare position of the stars across the panels in Figure 5). Across the economic dimension (expenditures), for example, we find that assuming compliance when in fact there is very little leads to greater expenditures. Recall by design, the *ad hoc* allocations have lower expenditures than the optimal policies because the central planner is not incurring the workability costs from deviating off of the allocation. At the same time, greater cumulative cases result when the opposite holds, that is, assuming no compliance when in fact there is compliance. We also see that in some instances that the combined effect of incorrectly assuming the wrong immunity and compliance can offset some deviations (e.g. see Figure 5 Panel C) while in other cases the results are dominated by non-compliance. Finally, when there is compliance to travel restrictions the *ad hoc* allocation performs worse than any of the optimal allocations, while the *ad hoc* allocation performs relatively well when there is no compliance to travel restrictions. Varying the level of scarcity does not change the qualitative nature of results (see figures A13 and A14 for when vaccine capacity is 5% and 15% respectively), except for one anomaly where the *ad hoc* does not always perform worse under assumptions on compliance to travel restrictions (Figure A14).

We also investigate the robustness of the optimal allocations when the demographic characteristics are heterogeneous across jurisdictions. When jurisdictions have a different case-fatality ratio, the *ad hoc* allocation performs better than the optimal allocations when considering cases as the main health outcome (Figure A15). However, this approach is misleading because when case-fatality ratios are heterogeneous across jurisdictions, the cumulative aggregate number of cases (all jurisdictions together) is a poor outcome measure as a case in one place is not the same as a case in another jurisdiction. In this setting, the disease burden and cumulative damages give a more accurate depiction of the situation. In fact, while the *ad hoc* allocation outperforms the optimal allocations in terms of cumulative cases, it performs considerably more poorly when considering cumulative damages. We generally find that the optimal allocations outperform the *ad hoc* allocation in all scenarios considered (Figure A16). When jurisdictions have a different contact rate, the same pattern as in Figure 5 holds in the sense that when there is compliance to travel restrictions, the optimal allocations outperform the *ad hoc* allocation, while the *ad hoc* generally performs better than the optimal allocations when there is noncompliance to travel restrictions (Figure A17).

### 3.4 Sensitivity Analyses

The previous section considers the robustness of optimal allocations to incorrect assumptions about parameters (e.g. assuming permanent immunity while in fact it is temporary). Public health officials will also want to know how much optimal allocations change when parameters change (e.g. because vaccine effectiveness is lower against a new strain of the virus). We address those questions in this section. While both sets of analyses address parameter uncertainty, you can consider in this section that the uncertainty is resolved before the public health officials have to make the vaccine allocation, while in the previous section the uncertainty was not resolved and public health officials had to choose allocations based on potentially incorrect assumptions.

Two key parameters in our analysis are the scale of the workability cost (*c*_*A*_ in Equation (9)) and the level of vaccine effectiveness (see Appendix C for more details). While imposing the *ad hoc* rule *ex ante* implicitly means that the cost of deviating from the *ad hoc* allocation is infinite, in practice it is likely finite but hard to quantify, as it depends on logistical, political, and cultural factors. We investigate the sensitivity of our results by solving for optimal vaccine allocation over a range of values. We find greater deviations off of the *ad hoc* at lower workability costs resulting in greater differences in cumulative cases, and smaller deviations as the workability cost parameter increases (Figure A18 panels A, B, C, and D). Specifically, we find that when the cost is in the neighborhood of the VSL (*c* in Equation (7) and Figure A18 black line represents the VSL), that the planner no longer deviates from the *ad hoc*.

The base case parameter for vaccine effectiveness we utilized in the paper is based on estimates of the influenza vaccine [51] (see Appendix A for more details) and represents a conservative estimate similar to the lower efficacy vaccines listed for emergency use listing by WHO (i.e. the Sinopharm COVID-19 vaccine, see e.g. [10]). Evidence from other COVID-19 vaccines (e.g. Pfizer/BioNTech and Moderna vaccines) suggest that effectiveness could be considerably higher than our base case scenario [11], but preliminary evidence suggests that vaccine efficacy is lower against new variants (e.g. the Delta variant, aka lineage B.1.617.2; see e.g. [12] and see [52] for a detailed report). We find that the more effective a vaccine is, the more a central planner would want to deviate from the *ad hoc* allocation (in blue; Figure A19 panels A, B, C, and D). As a result of this greater deviation, we see a larger difference in terms of the reduction in cumulative cases (in red; Figure A19 panels A, B, C, and D).

## 4 Conclusion

Recent studies have discussed how a vaccine against the coronavirus disease (COVID-19) should be allocated within a geographical area (see for instance [1; 2; 3]) and on a global scale (see for instance [4; 5; 6; 7]). Building off the spatial-dynamic literature in epidemiology, we contribute to this body of work by addressing the question of distributing a scarce allotment of COVID-19 vaccine across smaller geographic areas, such as counties or states, and by showing how optimal allocations perform when vaccine allocation must be determined before uncertainty about key behavioral (i.e. compliance to travel restrictions) and epidemiological (i.e. the length of immunity to the disease) factors are resolved. Countries receiving allotments of vaccines via COVAX—a World Health Organization (WHO) led initiative aimed to provide an equitable access to COVID-19 vaccines—could follow NASEM’s [8] and WHO’s [9] principle and allocate a COVID-19 vaccine to jurisdictions within their borders based on the jurisdictions’ population size. While this approach prioritizes an important principle—equity of distribution—we show in this paper that there are potential economic and public health benefits of deviating from such an allocation rule. The optimal allocation rule we derive is more closely aligned with another important principle—equity of outcomes— and this paper provides policymakers with information on the trade-offs involved with different allocation rules. We also highlight under which behavioral and epidemiological conditions the optimal allocation performs relatively better and when the *ad hoc* allocation is a good approximation of the optimal allocation. There are many factors that come into play in these allocation decisions and the methodology proposed here provides a way to benchmark these rules to illustrate the trade-offs. Other methodologies that do not solve for the optimal policies are left to benchmark one set of *ad hoc* rules against another, where the set of possible *ad hoc* rules is infinite.

We considered several different scenarios where the length of immunity, the compliance to travel restrictions, the size of the vaccine allotment, and the demographics across jurisdictions are varied. In most of these scenarios, we find that priority should be given to jurisdictions that initially have lower disease burden. The intuition behind this result—already put forward by Rowthorn et al. [20] when investigating optimal control of epidemics in a scenario where no immunity to the disease is developed—is that the priority should be to protect the greater population of susceptible individuals, and that focusing on a subset of the population, rather than on the entire population, can make a significant difference [53]. We find that a larger allotment of vaccine can lead to the opposite result for a short period of time at the beginning of the time horizon, and we find that the high burden jurisdiction should be prioritized when it has more essential workers, as long as its infection level is increasing and remains higher than the jurisdiction with fewer essential workers. Our results also suggest that deviations from an *ad hoc* allocation rule based on population size are highly beneficial when one jurisdiction has a population with higher mortality rates.

We also show the value of complying to a travel restriction, as compliance leads to lower cumulative damages across both jurisdictions, regardless of whether immunity is permanent or temporary. The reduction in cumulative damages is particularly important for the jurisdiction with fewer infected individuals. Considering nonlinear damages due to an overload of health care systems [54; 55] and a corresponding varying death rate due to scarce intensive care unit beds [24], and other second-order problems such as consumption losses [56; 57; 58], excess mortality [59], and psychological distress [60] could further highlight the benefits of complying to travel restrictions.

Despite having to pay a workability cost for deviating from the *ad hoc* allocation, we show that it is still in the interest of the central planning agency (e.g. the central government) to deviate from this rule of thumb for a wide range of values we considered; this result holds in all scenarios we considered in our analysis. We considered *ad hoc* allocation rules that favor “speed and workability” (put forward by NASEM [8], and WHO has a similar principle [9]). Other allocation rules are possible. For instance, in the base case of our paper, we assumed identical contact rates across jurisdictions. In turn, this implied that the movement within a given jurisdiction is assumed to be identical across jurisdictions. In practice, population mobility likely differs from one jurisdiction to another and an *ad hoc* allocation could be based on population mobility and contact structure. The methodology employed in this paper can investigate the trade-offs of other *ad hoc* rules and as a result, can offer potentially important information to policymakers that face the challenge of allocating scarce COVID-19 vaccines to their jurisdictions.

There are, however, important factors that have received significant attention in the literature that we should fully incorporate in future research. For example, the composition of the population of a jurisdiction is assumed to be homogeneous. To mimic the fact that the virus disproportionately affects elderly people [14] and/or people with pre-existing conditions [61], and also to mimic the fact that the risk of infection is highly occupation dependent [15], we simply assumed that the average case-fatality ratio and contact rate was higher in one jurisdiction. In practice, however, the composition of a population within a given jurisdiction is not homogeneous. Further research combining heterogeneity both across jurisdictions in the form of different disease burden and within jurisdictions in the form of different risk of complications and risk of infection could add additional valuable insights into the trade-offs inherent in these different allocations rules.

Finally, while our paper and most of the discussion revolves around the allocation of a vaccine, a similar allocation problem may arise if an antiviral drug were to become available (for a discussion on antiviral treatments for SARS-Cov-2, see [62]). Because drugs and vaccines have different goals—treating infected individuals and prophylaxis, respectively—the economic and public health trade-offs of different allocation rules may be unique to the type of pharmaceutical intervention. Future work considering the joint allocation question of antiviral drugs and vaccines could be valuable in understanding the trade-offs and complementarities between these different pharmaceutical interventions.

## Data Availability

The parameter values used to run the numerical simulations are publicly available from the citations included in the text.

https://github.com/fmcastonguay/SpatialAllocationCOVID19

## Appendices

### A Parameterization

#### A.1 Epidemiological Model

According to Diekmann et al. [1], the basic reproduction ratio *R*_0_ of any disease is given by the expected number of secondary infection caused a by a typical infected individual over its entire infectious period, at a disease-free equilibrium. In the most basic epidemiological model, the *R*_0_ is simply given by the contact rate multiplied by the mean infectious period. When considering more complex models—as the two-jurisdiction SEIR model in this paper—one needs to use the next-generation matrix and find its dominant eigenvalue to find the *R*_0_ [1]. Denote two matrices by *F* and *V*, and let the *ij*^th^ element in *F* represents the rate at which infected individuals in population *j* produce new infections in population *i*, and the *ij*^th^ element in *V* represents the transition rate between (*i* ≠ *j*), or out of (*i* = *j*), infectious compartments [2]; the next-generation matrix is equal to *-FV* ^*-*1^. In the model presented in this paper,

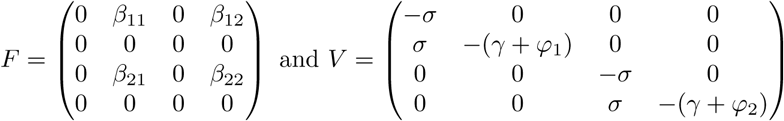

where the four rows of *F* and *V* refer to the *E*_1_, *I*_1_, *E*_2_ and *I*_2_ equations, respectively. Note that both matrices *F* and *V* are derived under the assumption of introducing a single exposed individuals in an otherwise susceptible population (for more details on how to construct the next-generation matrix in a SEIR model, see [3]). Given we assume that *β*_11_ = *β*_22_ = *β*_*ii*_ and *β*_12_ = *β*_21_ = *β*_*ij*_, and when in our main analysis we let *φ*_1_ = *φ*_2_ = *φ*, the basic reproduction ratio of our model simplifies to,

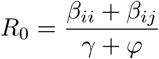

for *i* = 1, 2, *j* = 1, 2, and *i* ≠ *j*. We set the basic reproduction ratio *R*_0_ = 1.43, according to estimates of the *R*_0_ from Li et al. [4] and using estimates of the effect of nonpharmaceutical interventions on the *R*_0_ from Tian et al. [5]. We assume a mean recovery period 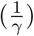 of 5 days [6], and a case-fatality ratio of 1.78% (adjusted for misreporting, see [7]) to calibrate the rate of disease induced mortality, *φ*. Parameters *β*_*ii*_ and *β*_*ij*_ are then calibrated assuming what Tian et al. [5] call a “medium effect of the [nonpharmaceutical] control” when there is compliance to travel restrictions, and a “lower effect of the [nonpharmaceutical] control” when there is no compliance to travel restrictions (for evidence of structural changes in mobility following the COVID-19 lockdown, see [8]); this yields *R*_0_ *≈* 1.4 with compliance to travel restrictions, and *R*_0_ *≈* 2.1 when there is no compliance to travel restrictions. The mean latency period 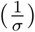, which one needs to know to calculate matrix *V* even though it does not appear in the basic reproduction ratio, is assumed to last 3 days [6].

#### A.2 Economic Model

To quantify damages, we use the value of statistical life recommended by the Environmental Protection Agency.^1^ The disability weight^2^ associated with COVID-19 infection is assumed to be equivalent to a lower respiratory tract infection, which is a disability weight of *w* = 0.133 on a scale from zero (perfect health) to one (death).^3^ This disability weight thus allows for a comparison between the individuals that are infected with the disease but do not die, and the individuals that die from its complications.

Expenditures related to the pharmaceutical intervention are based off estimates of vaccine costs. Numerous governments around the world, including the U.S. federal government, have contracted biotech companies producing COVID-19 vaccines; governments pay money in exchange of a guaranteed number of doses of COVID-19 vaccines. These estimates and the prices of current influenza vaccine turn out to be approximately 20 U.S. dollars per dose, with two doses per individual; this is the value we chose in our analysis.^4^

The value of the workability cost^5^ is based on a certain proportion of the value of statistical life; in the base case, we assume it to be 3 orders of magnitude smaller. All costs in the model are assumed to be discounted at a 1.5% annual rate (see [12] for a discussion about discounting health-related expenditures).

#### A.3 Parameter Levels

Table A1 below summarizes the main set of parameter values we used in the numerical simulation.

**Table A1:**
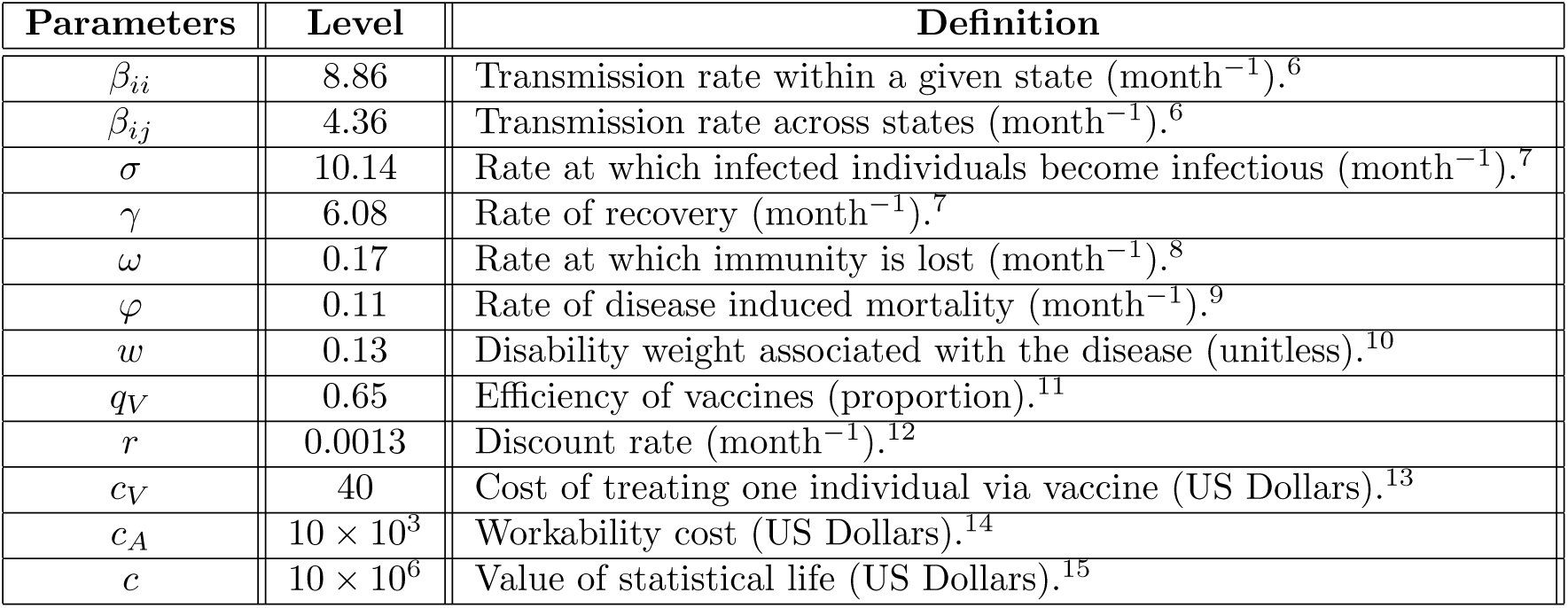
Parameter levels used in the numerical simulation.

### B Optimization

#### B.1 Boundary Conditions

To yield the initial conditions of the optimal control problem, we calibrated the model using the above parameter values and simulated out a COVID-19 outbreak in two identical jurisdictions, where we assumed there was one exposed individual in an otherwise entirely susceptible population of 10 million individuals. We assumed that both jurisdictions undertook nonpharmaceutical interventions that had a “medium effect” on the basic reproduction ratio [5] (i.e. that there was perfect compliance to travel restrictions). After simulating out the disease dynamics for a period of eight months and two weeks, and eight months and three weeks for Jurisdiction 1 and Jurisdiction 2 respectively, the initial conditions yield were:

**Table A2:** Initial conditions of the numerical simulation.

| Jurisdiction | $N_i$ | $S_i$ | $E_i$ | $I_i$ | $R_i$ |
| --- | --- | --- | --- | --- | --- |
| Jurisdiction 1 | 1 | 0.9074 | 0.0103 | 0.0143 | 0.0667 |
| Jurisdiction 2 | 1 | 0.8662 | 0.0138 | 0.0196 | 0.0986 |

We assume that the terminal conditions (i.e. the conditions on state variables in *t* = *T*, the final time period) are free to be optimally determined. Formally, the initial and terminal conditions of the ten state variables are such that:

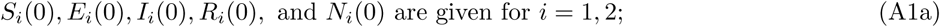

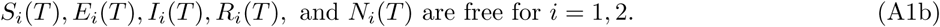

#### B.2 Nonnegativity and Upper-Bound Constraints

State variables *S*_*i*_, *E*_*i*_, *I*_*i*_, *R*_*i*_, and *N*_*i*_ for *i* = 1, 2 are subject to nonnegativity and physical constraints. Formally:

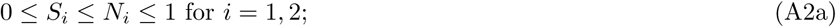

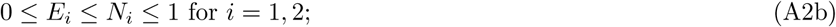

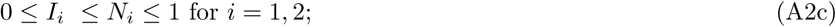

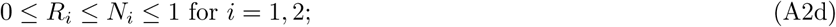

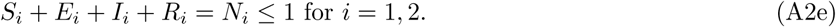

Control variables are modelled as direct controls (see examples in [17; 18; 19]) and can be interpreted as a reduction in the number of susceptible individual in a given time period (i.e. a month). Formally, the constraints on the control variables are given by:

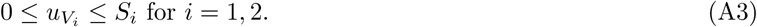

Because of a limited supply of vaccines (see details below), the physical upper-bound on constraints (A3) will only be binding when capacity constraint is nonbinding. When this occurs, it means that there are fewer susceptible individuals than there are available vaccines.

#### B.3 Capacity Constraints of the Pharmaceutical Interventions

For completeness, we also include the capacity constraints already mentioned in the main paper. In addition to the physical constraints on the control variables, the aim of our paper is to study how to allocate a limited allotment of vaccines. Hence, the control variables are also subject to

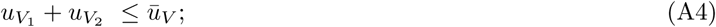

when the central planning agency decides to potentially deviate from the *ad hoc* allocation of vaccine. Conversely, the *ad hoc* constraints are:

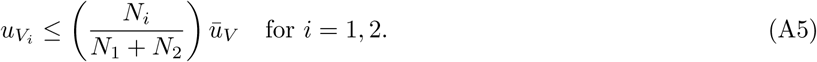

As mentioned in the main paper, the total available quantity of vaccine (*ū*_*V*_) represents a certain percentage (5%, 10%, or 15%) of the total population size.

#### B.4 Numerical Methods

Pseudospectral collocation approximates the continuous time optimal control model with a constrained nonlinear programming problem (see [20; 21; 22; 23] for other applications of this technique). The dynamic controls to our problem—i.e. the vaccine allocation—are approximated by a polynomial of degree *n* (determined by the number of collocation points) over a period from *t* = 0 (date at which the vaccine starts to be administered) to *t* = *T* (assumed to be four months after the vaccine administration) [24]. The residual error of the constraints is minimized by the algorithm at the *n* collocation points, where *n* is chosen to have a reasonable speed of convergence to a solution and a low numerical error. Here, we chose 60 collocations points. In this sort of problem, the main advantage of this approach over more usual methods to solve such two-point boundary problems, such as shooting methods, is that nonnegativity constraints (e.g. on the number of infected individuals) and upper-bound constraints (mimicking e.g. vaccine capacity constraints) on state and control variables can be directly incorporated in the problem [25]. This method thus allows us to find optimal solutions that may lay on the boundary of the control set for a certain period of time. For COVID-19 vaccines, this is likely due to the scarcity of the supply of vaccine in the short-term. Another advantage of this method is the ability to deal with large-scale dynamical systems, such as the one presented here with ten state variables and two control variables. The solution was found using TOMLAB (v. 8.4) [26; 27] and the accompanying PROPT toolbox [28]. The approximate nonlinear programming problem is solved using general-purpose nonlinear optimization packages (e.g. KNITRO, SNOPT and NPSOL).

### C Figures

#### C.1 Base Case: Homogeneous Demographic Characteristics

##### C.1.1 Compliance and Noncompliance to the Travel Restrictions

**Figure A1:**
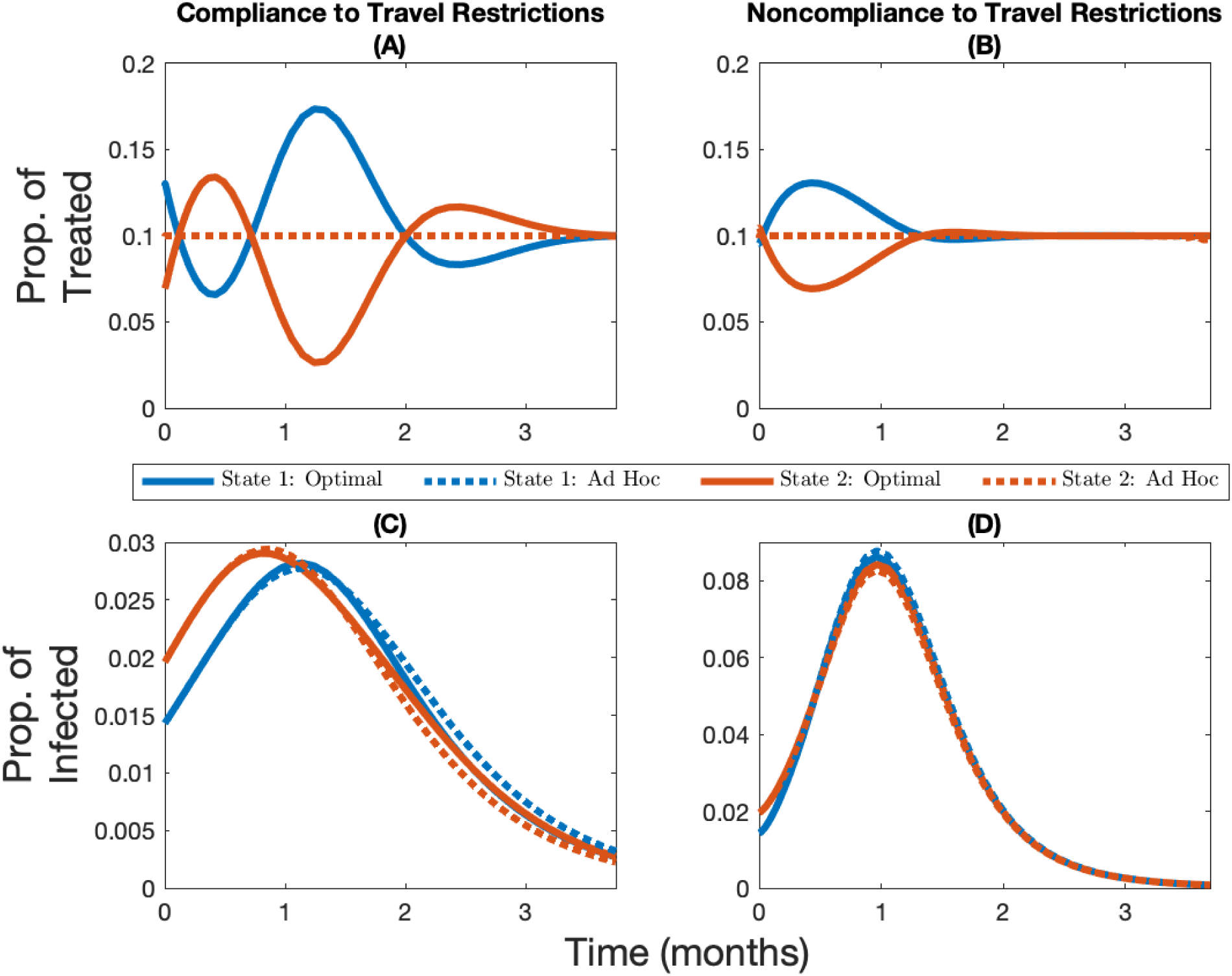
Permanent immunity with and without compliance to travel restrictions. Change over time in the optimal and *ad hoc* allocations (panels A and B) and the corresponding infection levels (panels C and D) for State 1 (in blue, the initially lowest-burdened state) and State 2 (in red, the initially highest-burdened state) depending on whether there is compliance to travel restrictions (panels A and C) or not (panels B and D) for the case where the vaccine capacity constraint is 5% and immunity lasts six months.

##### C.1.2 Vaccine Capacity Constraints when Immunity is Permanent

**Figure A2:**
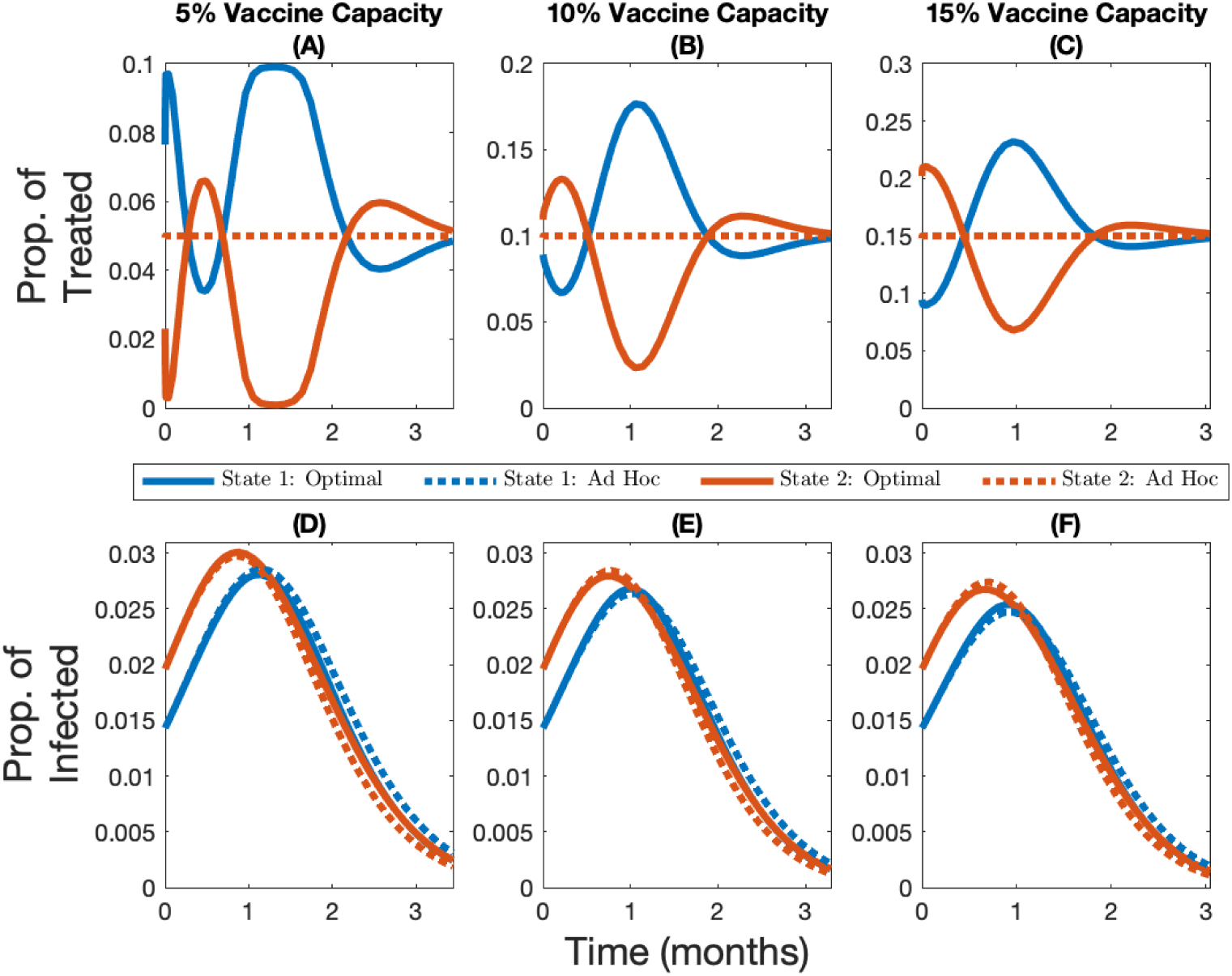
Permanent immunity and compliance to travel restrictions with 5%, 10%, and 15% vaccine capacity. Change over time in the optimal and *ad hoc* allocations (panels A, B, and C) and the corresponding infection levels (panels D, E, and F) for State 1 (in blue, the initially lowest-burdened state) and State 2 (in red, the initially highest-burdened state) depending on whether capacity is 5% (panels A and D), 10% (panels B and E), or 15% (panels C and F), for the case where immunity is permanent and there is compliance to travel restrictions.

##### C.1.3 Vaccine Capacity Constraints when Immunity is Temporary

**Figure A3:**
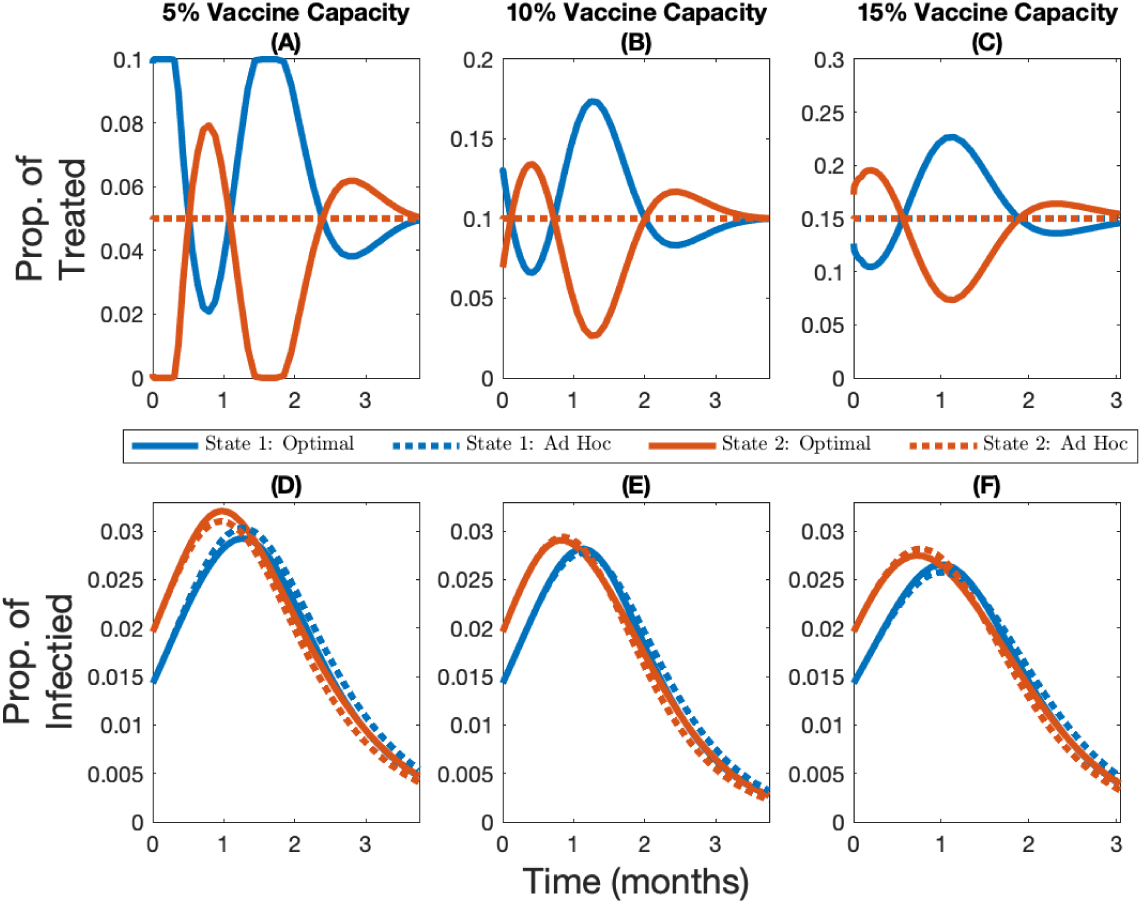
Temporary immunity and compliance to travel restrictions with 5%, 10%, and 15% vaccine capacity. Change over time in the optimal and *ad hoc* allocations (panels A, B, and C) and the corresponding infection levels (panels D, E, and F) for State 1 (in blue, the initially lowest-burdened state) and State 2 (in red, the initially highest-burdened state) depending on whether capacity is 5% (panels A and D), 10% (panels B and E), or 15% (panels C and F), for the case where immunity lasts six months and there is compliance to travel restrictions.

**Figure A4:**
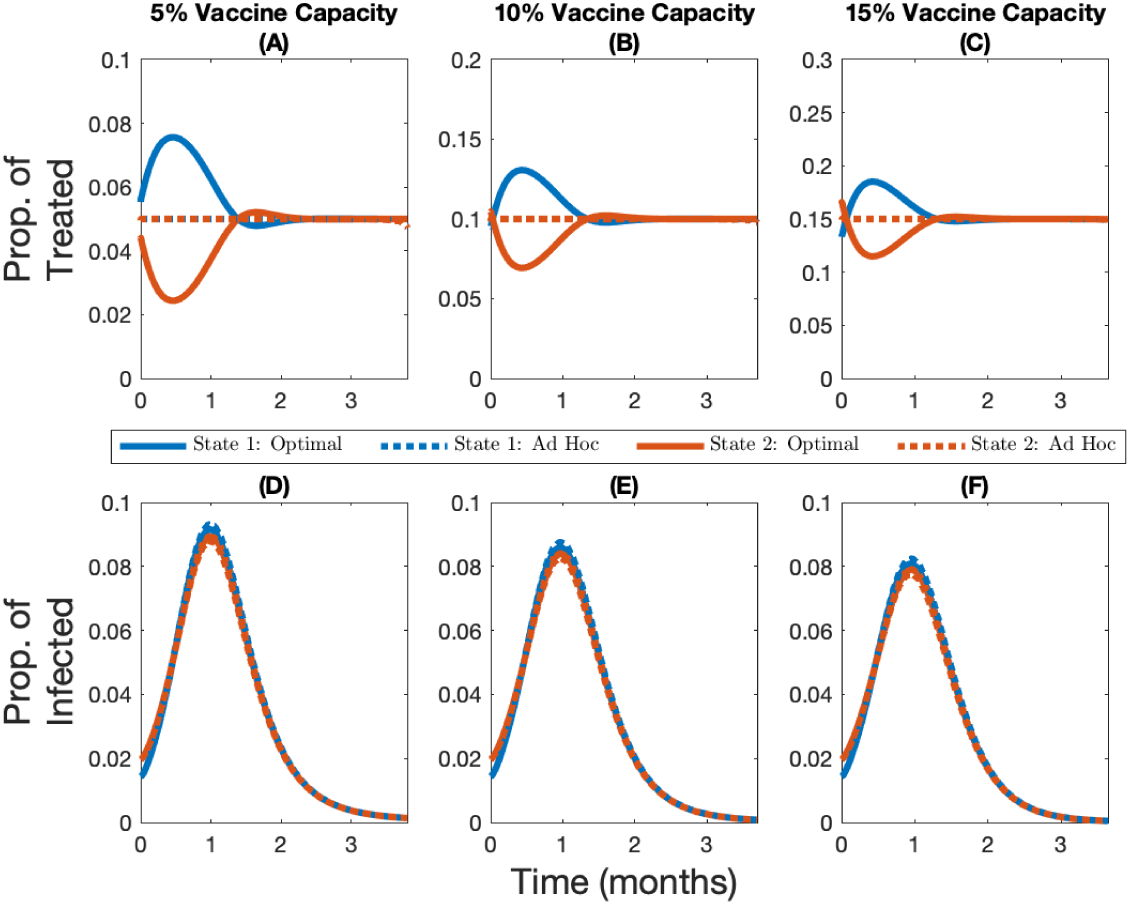
Temporary immunity and noncompliance to travel restrictions with 5%, 10%, and 15% vaccine capacity. Change over time in the optimal and *ad hoc* allocations (panels A, B, and C) and the corresponding infection levels (panels D, E, and F) for State 1 (in blue, the initially lowest-burdened state) and State 2 (in red, the initially highest-burdened state) depending on whether capacity is 5% (panels A and D), 10% (panels B and E), 15% (panels C and F), for the case where immunity lasts six months and there is no compliance to travel restrictions.

##### C.1.4 Permanent vs Temporary Immunity

**Figure A5:**
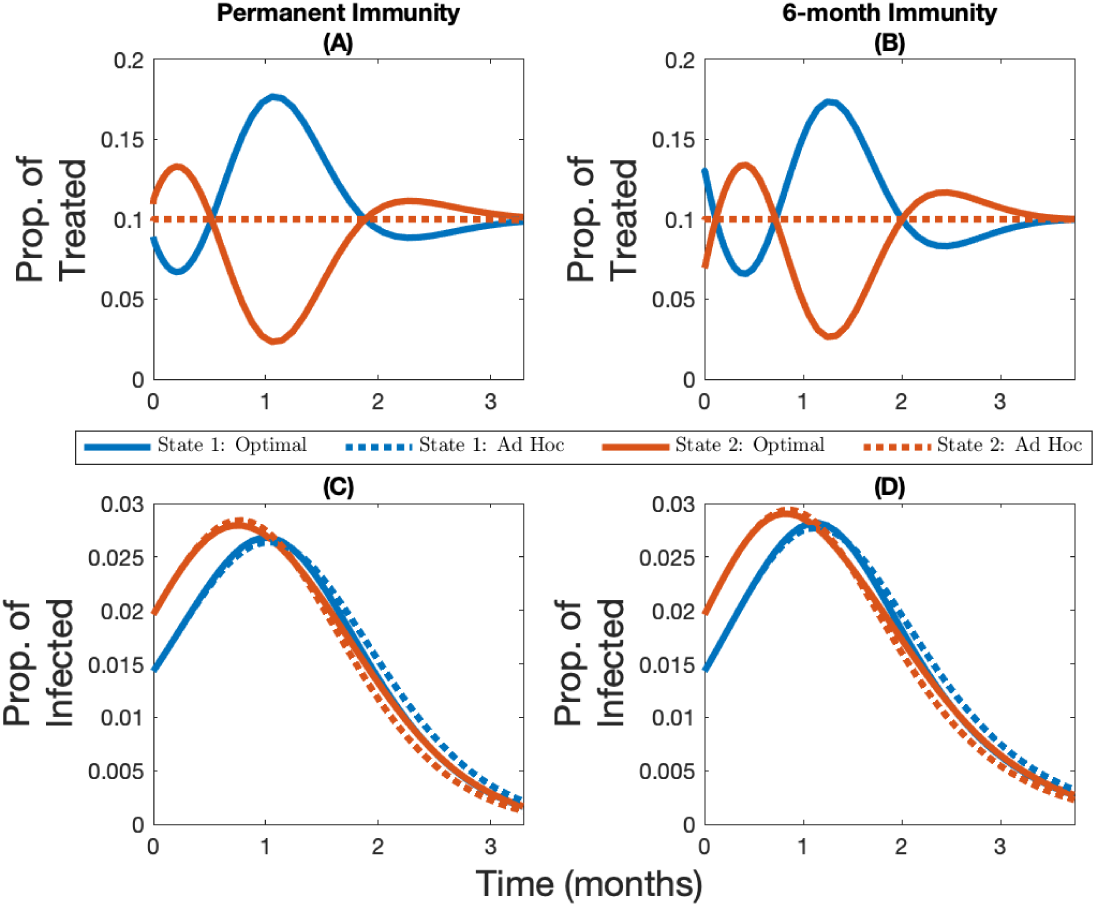
Compliance to travel restrictions with permanent and temporary immunity. Change over time in the optimal and *ad hoc* allocations (panels A and B) and the corresponding infection levels (panels C and D) for State 1 (in blue, the initially lowest-burdened state) and State 2 (in red, the initially highest-burdened state) depending on whether immunity is permanent (panels A and C) or lasts six months (panels B and D) for the case where the vaccine capacity constraint is 10% and there is compliance to travel restrictions.

**Figure A6:**
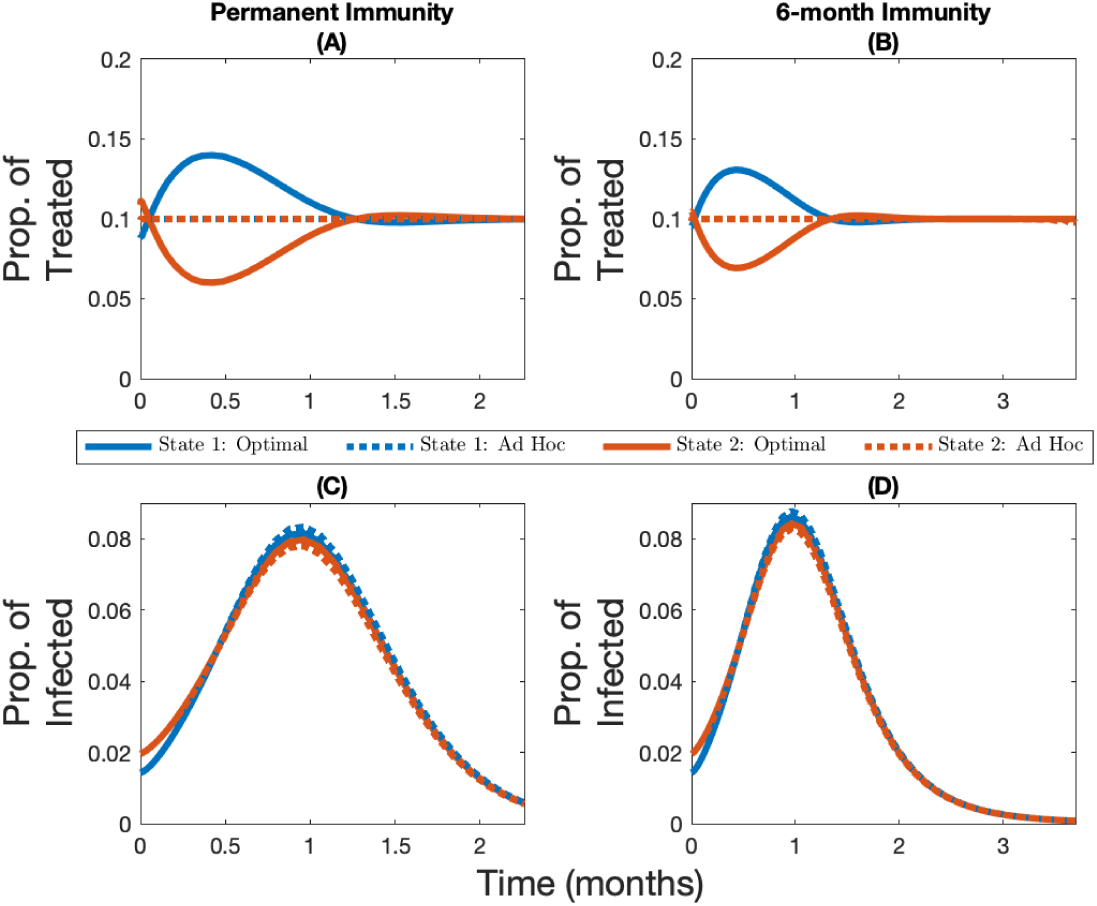
Noncompliance to travel restrictions with permanent and temporary immunity. Change over time in the optimal and *ad hoc* allocations (panels A and B) and the corresponding infection levels (panels C and D) for State 1 (in blue, the initially lowest-burdened state) and State 2 (in red, the initially highest-burdened state) depending on whether immunity is permanent (panels A and C) or lasts six months (panels B and D) for the case where the vaccine capacity constraint is 10% and there is no compliance to travel restrictions.

##### C.1.5 Cumulative Infection Levels

**Figure A7:**
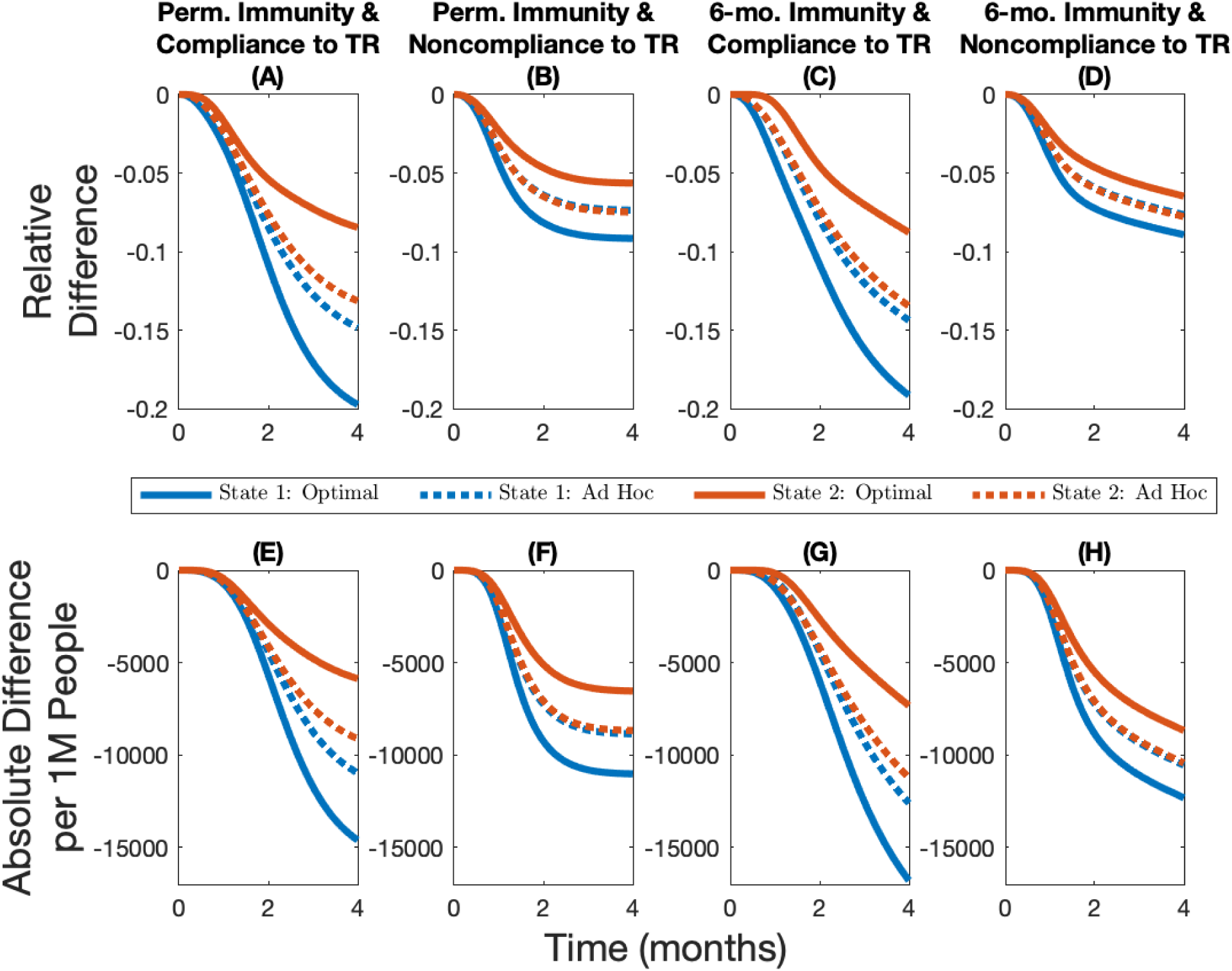
Epidemiological outcomes under different scenarios with a low vaccine supply. Cumulative relative difference (panels A, B, C, and D) and cumulative absolute difference per 1M people (panels E, F, G, and H) between the number of infections in different allocations rules and the no-vaccine case for different immunity–travel restrictions scenarios and for when vaccine capacity is 5%.

**Figure A8:**
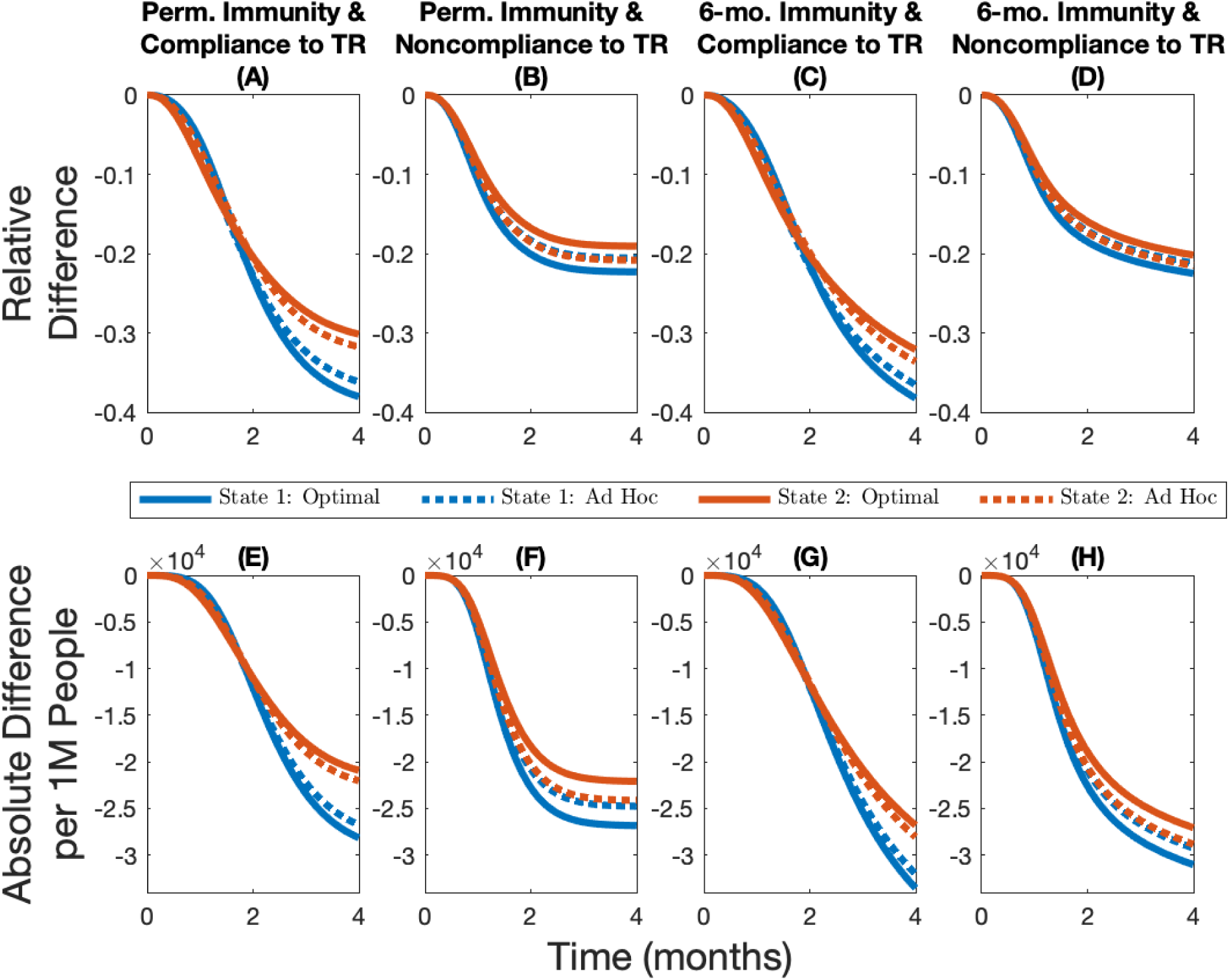
Epidemiological outcomes under different scenarios with a high vaccine supply. Cumulative relative difference (panels A, B, C, and D) and cumulative absolute difference per 1M people (panels E, F, G, and H) between the number of infections in different allocations rules and the no-vaccine case for different immunity–travel restrictions scenarios and for when vaccine capacity is 15%.

#### C.2 Heterogeneous Demographic Characteristics

##### C.2.1 Heterogeneous Case-Fatality Ratio

**Figure A9:**
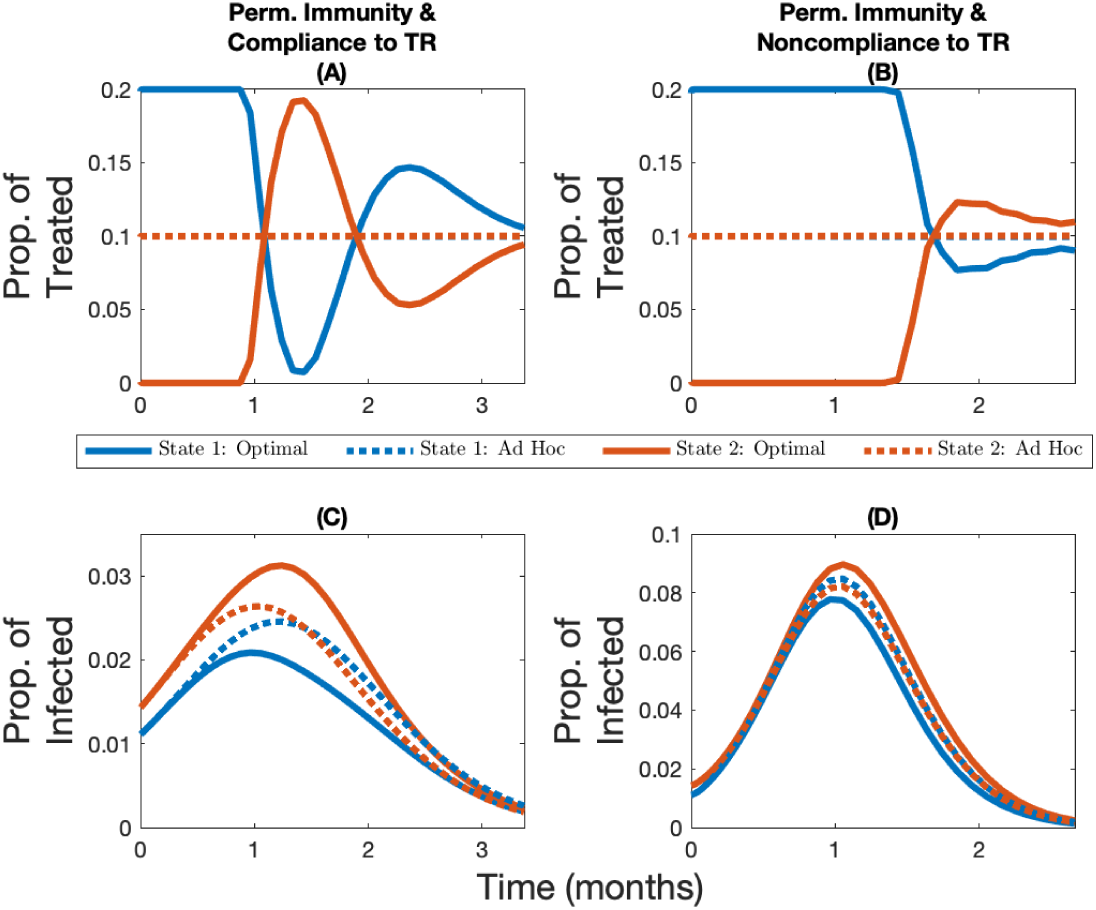
Vaccine allocation with and without compliance to travel restrictions. Change over time in the optimal and *ad hoc* allocations (panels A and B) and the corresponding infection levels (panels C and D) for State 1 (in blue, the initially lowest-burdened state) and State 2 (in red, the initially highest-burdened state) depending on whether there is compliance to travel restrictions (panels A and C) or not (panels B and D) for the case where the vaccine capacity constraint is 10%, immunity is permanent, and where the heterogeneity in the system comes from a varying case-fatality ratio (State 1 has a case-fatality ratio 1% higher than State 2).

**Figure A10:**
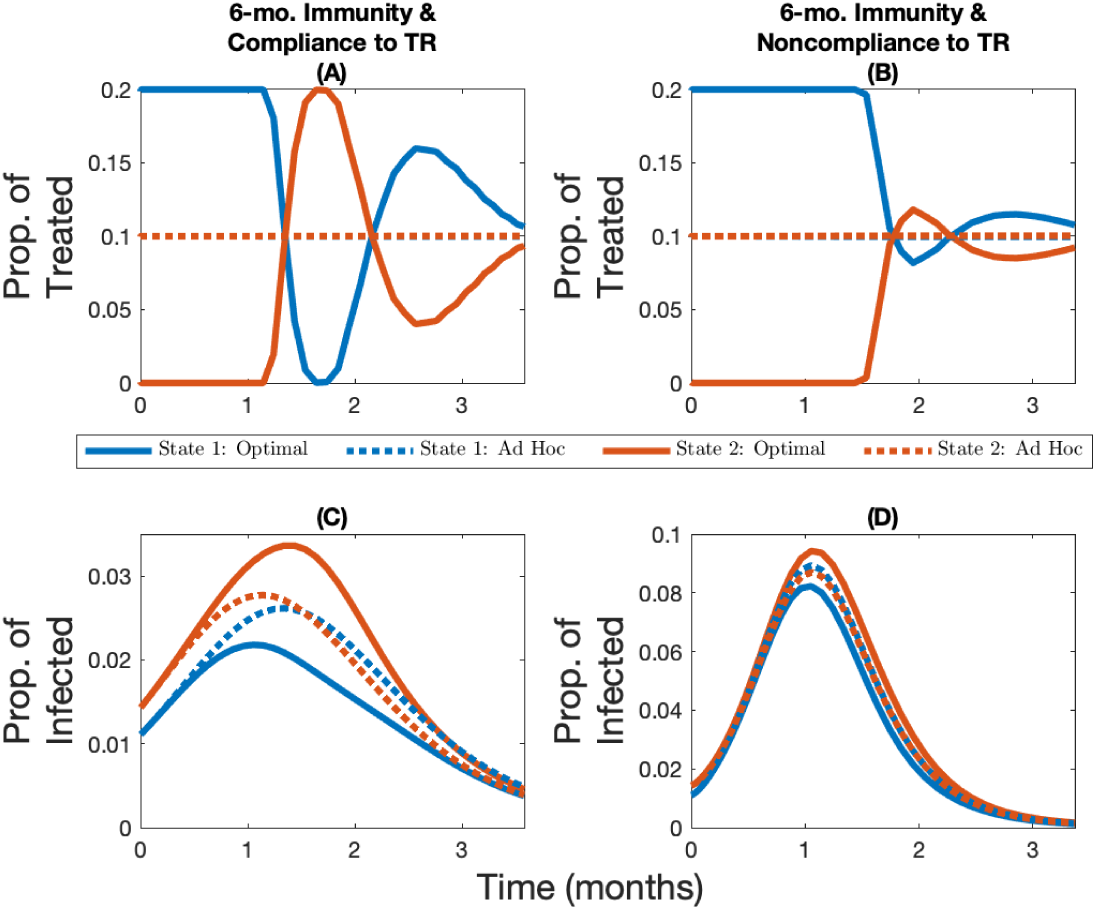
Vaccine allocation with and without compliance to travel restrictions. Change over time in the optimal and *ad hoc* allocations (panels A and B) and the corresponding infection levels (panels C and D) for State 1 (in blue, the initially lowest-burdened state) and State 2 (in red, the initially highest-burdened state) depending on whether there is compliance to travel restrictions (panels A and C) or not (panels B and D) for the case where the vaccine capacity constraint is 10%, immunity lasts six months, and where the heterogeneity in the system comes from a varying case-fatality ratio (State 1 has a case-fatality ratio 1% higher than State 2).

##### C.2.2 Heterogeneous Contact Rate

**Figure A11:**
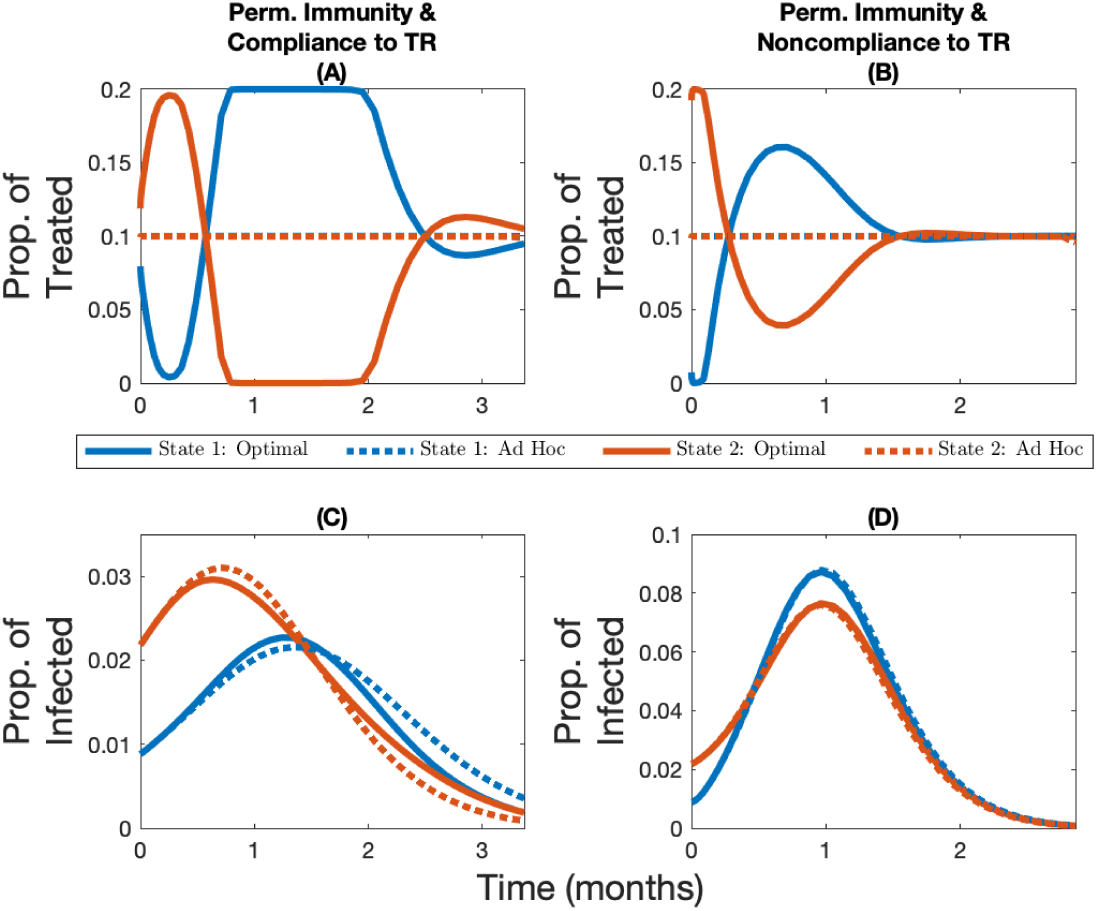
Vaccine allocation with and without compliance to travel restrictions. Change over time in the optimal and *ad hoc* allocations (panels A and B) and the corresponding infection levels (panels C and D) for State 1 (in blue, the initially lowest-burdened state) and State 2 (in red, the initially highest-burdened state) depending on whether there is compliance to travel restrictions (panels A and C) or not (panels B and D) for the case where the vaccine capacity constraint is 10%, immunity is permanent, and where the heterogeneity in the system comes from a varying contact rate (State 2 has a higher contact rate than State 1).

**Figure A12:**
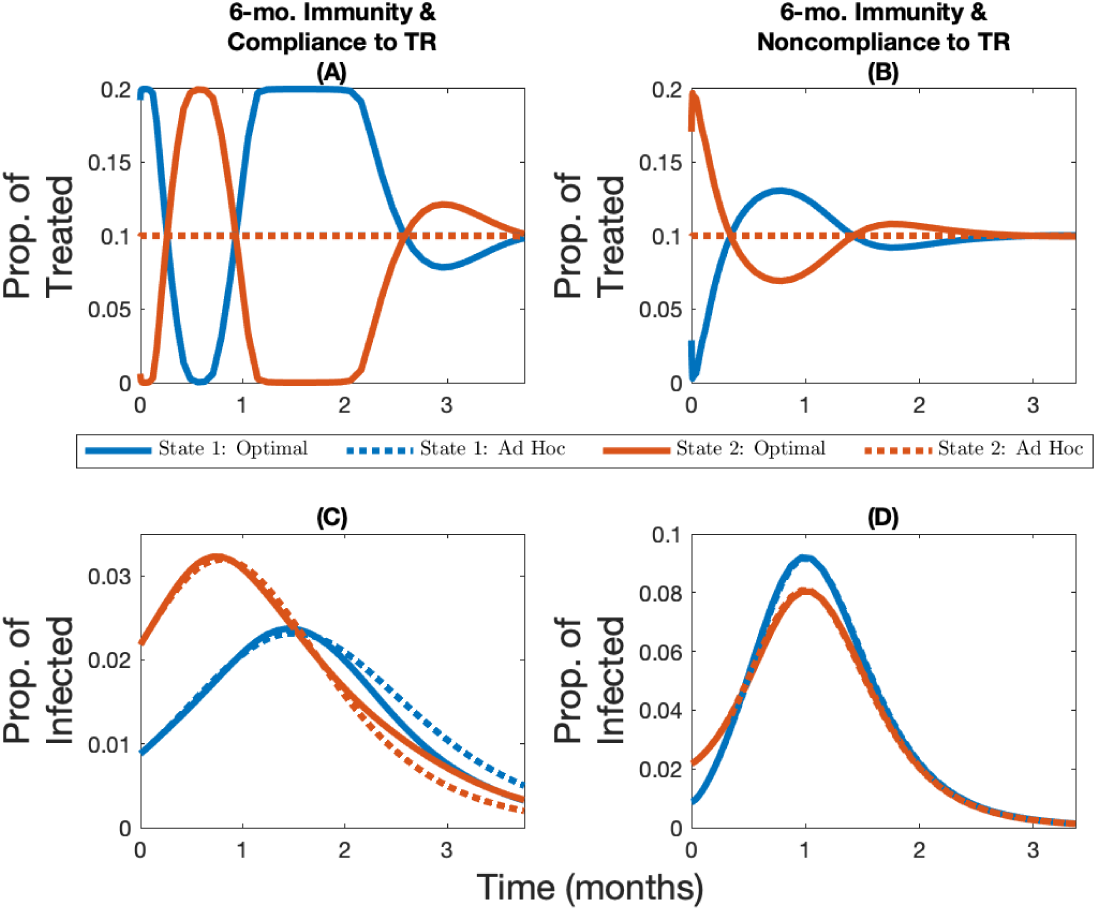
Vaccine allocation with and without compliance to travel restrictions. Change over time in the optimal and *ad hoc* allocations (panels A and B) and the corresponding infection levels (panels C and D) for State 1 (in blue, the initially lowest-burdened state) and State 2 (in red, the initially highest-burdened state) depending on whether there is compliance to travel restrictions (panels A and C) or not (panels B and D) for the case where the vaccine capacity constraint is 10%, immunity lasts six months, and where the heterogeneity in the system comes from a varying contact rate (State 2 has a higher contact rate than State 1).

#### C.3 Robustness of Spatial allocations

##### C.3.1 Base Case: Homogeneous Demographic Characteristics

**Figure A13:**
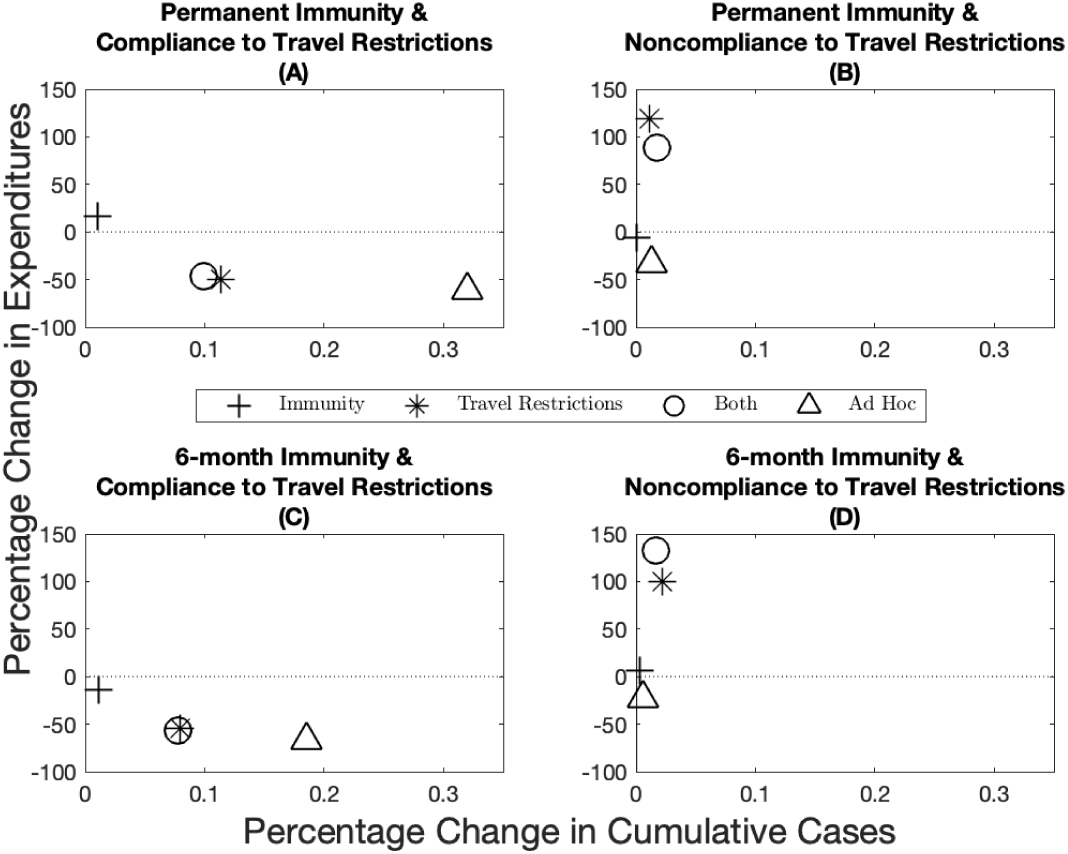
Robustness of epidemiological and economic outcomes under different scenarios. Percentage change in expenditures (y-axis) and percentage change in cumulative cases (x-axis) from the optimal allocation for different immunity–travel restrictions scenarios and for when vaccine capacity is 5%.

**Figure A14:**
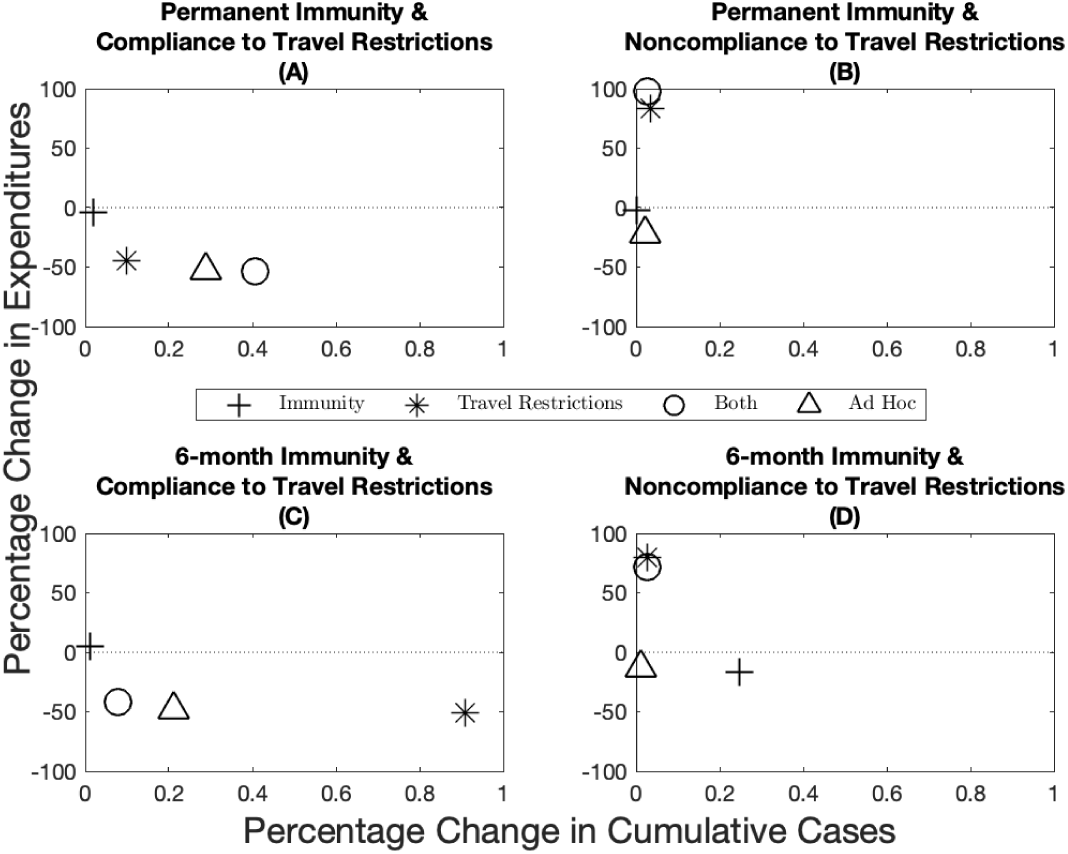
Robustness of epidemiological and economic outcomes under different scenarios. Percentage change in expenditures (y-axis) and percentage change in cumulative cases (x-axis) from the optimal allocation for different immunity–travel restrictions scenarios and for when vaccine capacity is 15%.

##### C.3.2 Heterogeneous Case-Fatality Ratio

**Figure A15:**
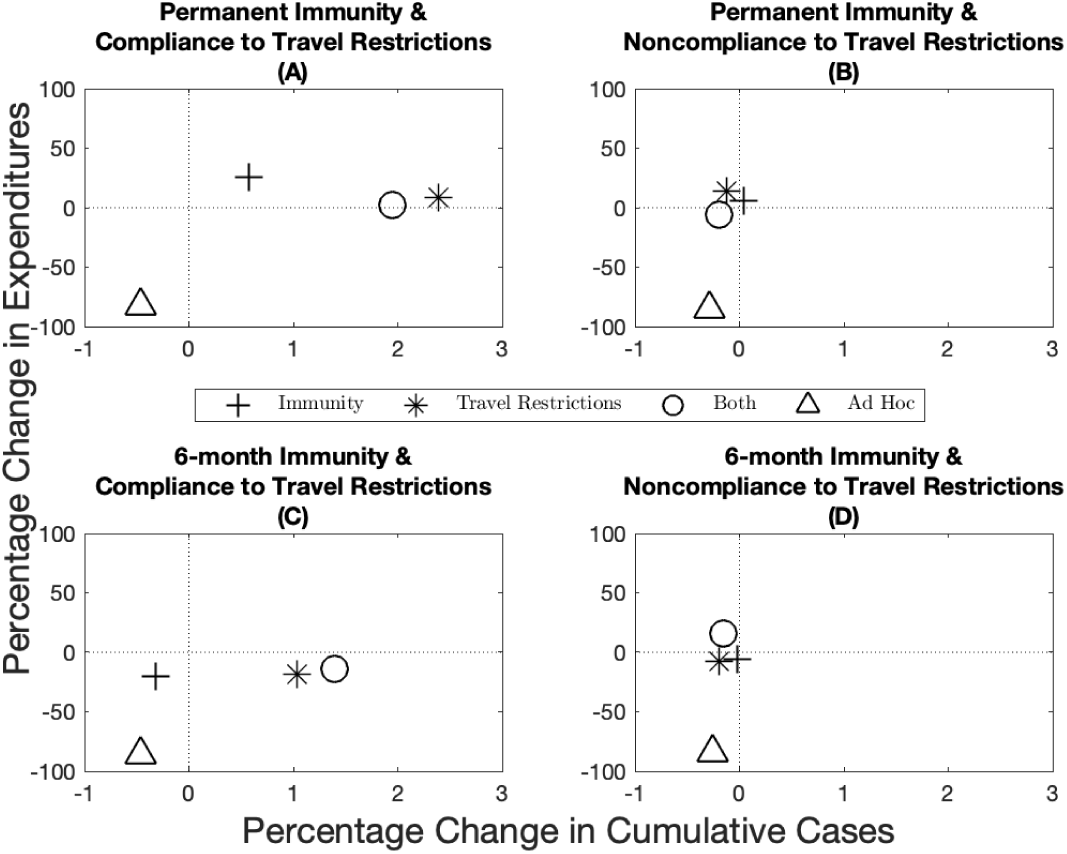
Robustness of epidemiological and economic outcomes under different scenarios when the source of heterogeneity is the case-fatality ratio. Percentage change in expenditures (*y*-axis) and percentage change in cumulative cases (*x*-axis) from the optimal allocation for different immunity–travel restrictions scenarios and for when vaccine capacity is 10%.

**Figure A16:**
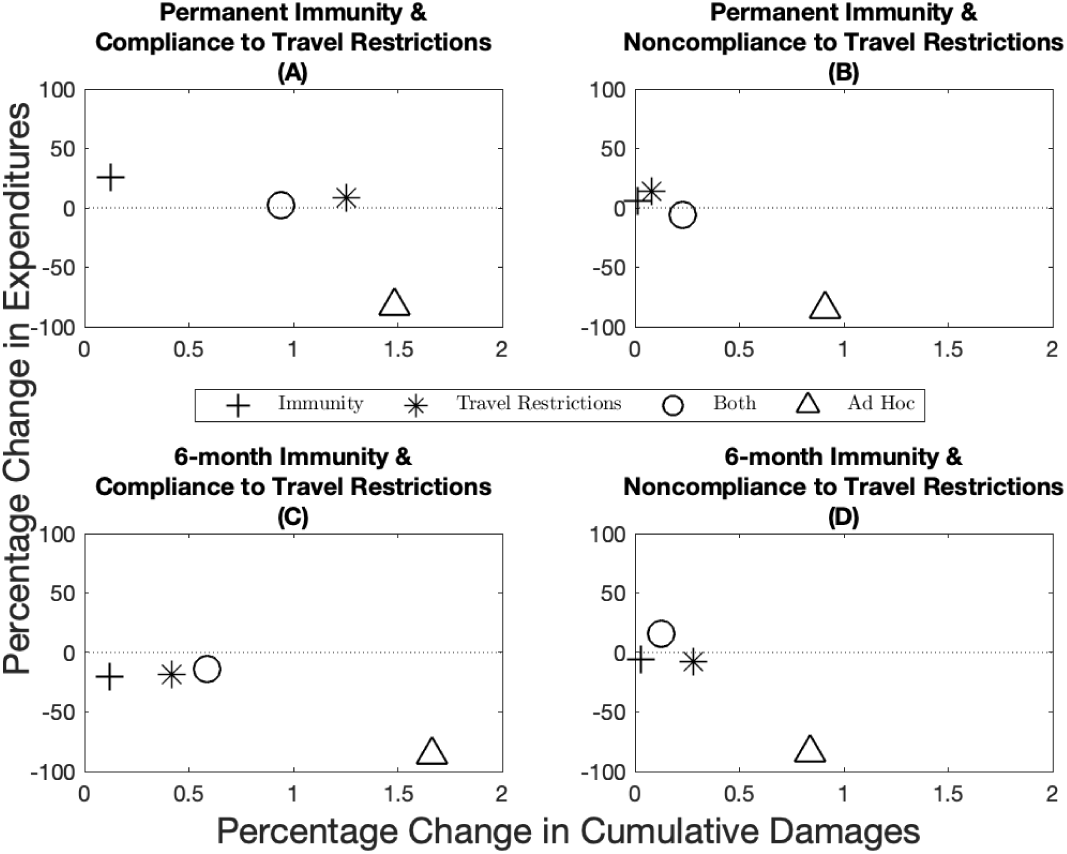
Robustness of epidemiological and economic outcomes under different scenarios when the source of heterogeneity is the case-fatality ratio. Percentage change in expenditures (*y*-axis) and percentage change in cumulative damages (*x*-axis) from the optimal allocation for different immunity–travel restrictions scenarios and for when vaccine capacity is 10%. Note that compared to Figure A15, the use of cumulative damages in this figure gives a more accurate depiction of the situation because cases across jurisdictions are not homogeneous when the case-fatality ratio is different.

##### C.3.3 Heterogeneous Contact Rate

**Figure A17:**
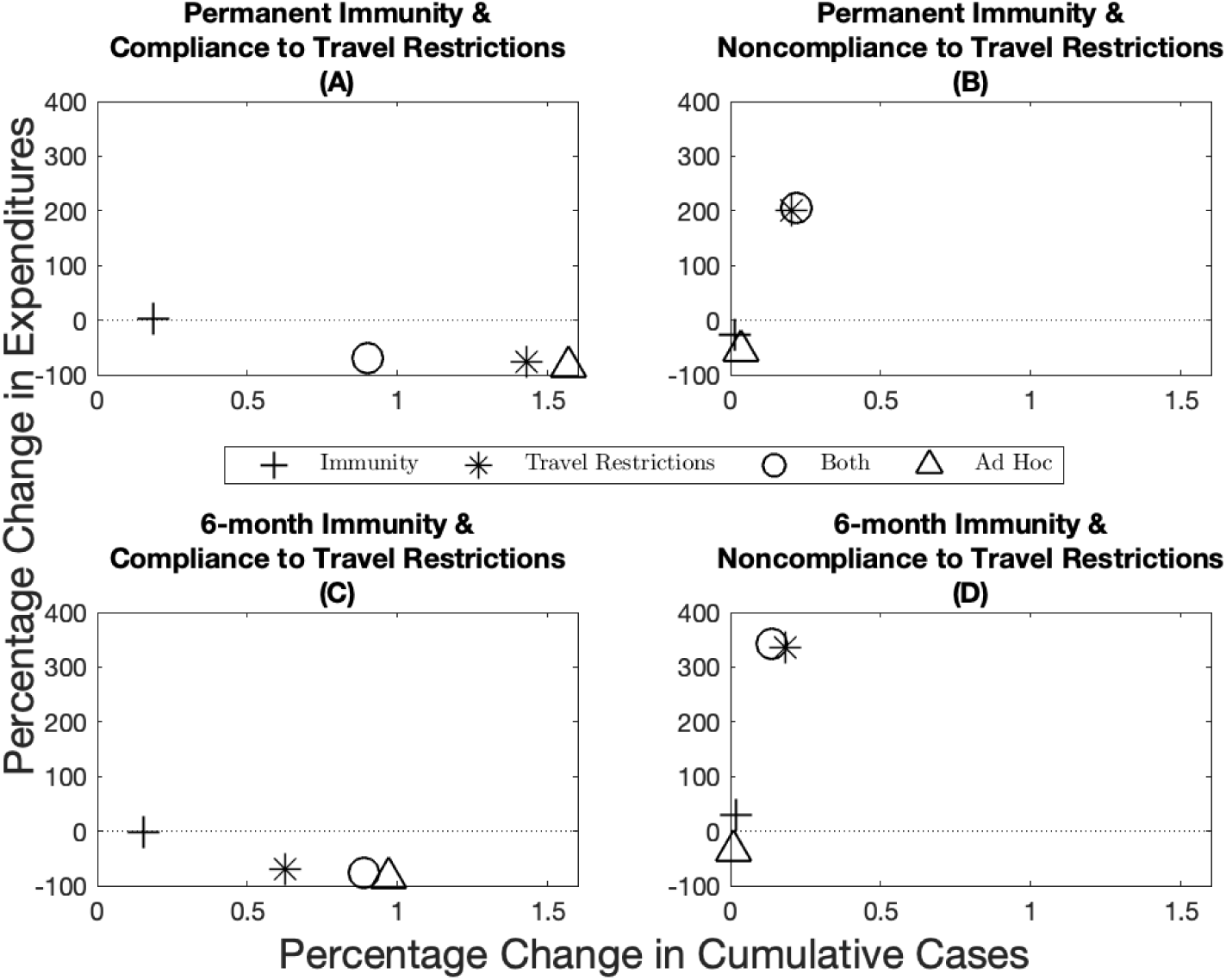
Robustness of epidemiological and economic outcomes under different scenarios when the source of heterogeneity is the contact rate. Percentage change in expenditures (*y*-axis) and percentage change in cumulative cases (*x*-axis) from the optimal allocation for different immunity–travel restrictions scenarios and for when vaccine capacity is 10%.

#### C.4 Sensitivity Analyses

##### C.4.1 Workability Cost

As mentioned above, imposing the *ad hoc* rule *ex ante* implicitly means that the central planning agency is essentially assuming that the cost of deviating from the *ad hoc* allocation is infinite. In practice, the workability cost is hard to quantify, because it depends on logistical, political, and cultural factors. It does however seem reasonable to assume, as we did in the paper, that the cost is finite. We investigate the sensitivity of our results by solving for the optimal vaccine allocation over time with levels lower and higher than the base case parameter in the paper. We summarize these results by plotting the variance^16^ of the optimal deviation in each time period from the *ad hoc* vaccine allocation (in blue; Figure A18 panels A, B, C, and D), and the difference in cumulative cases between the optimal and *ad hoc* allocation (in red; Figure A18 panels A, B, C, and D) as we vary the scale of the workability cost. Mathematically, as the workability cost approaches zero, the optimal control problem becomes linear in the controls, which implies that there is no adjustment cost associated with changing the allocation. Often times this can lead to extreme solutions (allocation goes to one state for a time period and then the other state, and so on).

Given the behavior and nature of the problem, therefore, we expect that at lower values of the workability cost parameter we will find higher variance of the deviation. This, in turn results in a higher performance of the optimal allocation relative to the *ad hoc* in terms of reduction in cumulative cases. When we increase the workability cost parameter, the cost parameter will eventually be on the same magnitude as the VSL (Figure A18 black line represents the VSL). When we reach levels this high, the optimal allocation converges towards the *ad hoc* and any differences in cumulative cases disappear.

We also show how amount of funds allocated to the workability cost over time compare to expenditures on the total vaccine cost (Figure A18 panels E, F, G, and H). If this ratio exceeds one, the planner is spending on aggregate more to deviate from the *ad hoc* than on treatments. These panels show that at low levels of the workability cost parameter, the total workability costs are small relative to the total vaccine costs. As the workability cost parameter increases, however, the total workability costs become more and more important relative to the total vaccine cost. Eventually, these costs begin to dominate the planners objective and the deviation between the *ad hoc* and optimal goes to zero.

**Figure A18:**
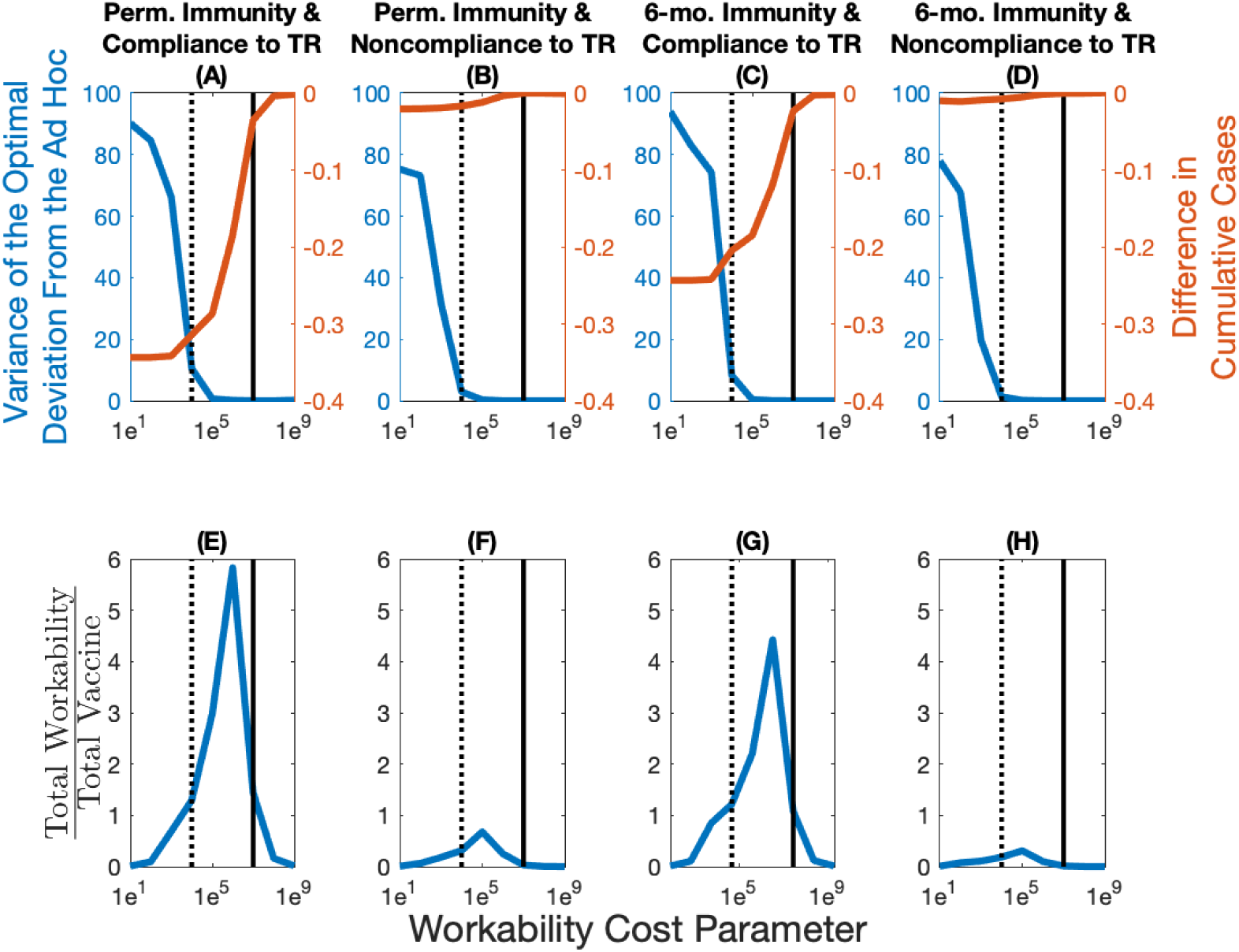
Sensitivity of optimal allocations and epidemiological and economic outcomes when varying the workability cost parameter. The variance of the optimal deviation in percentage (in blue; panels A, B, C, and D) represents an aggregate measure of the optimal deviation from the *ad hoc* allocation. The difference in cumulative cases between the optimal and *ad hoc* allocations (in red; panels A, B, C, and D) represents in percentage terms how well the optimal allocation outperforms the *ad hoc* allocation. The total workability cost over the total vaccine cost (panels E, F, G, and H) represents how many times more the total workability costs are relative to the total vaccine costs. The dotted vertical line represents the base case value of the workability cost parameter (1*e*^4^), while the full vertical line represents the value of statistical life (1*e*^7^).

##### C.4.2 Vaccine Effectiveness

The base case parameter for vaccine effectiveness we utilized in the paper is based on estimates of the influenza vaccine [15]; see Appendix A for more details. Recent evidence from the COVID-19 vaccines suggest that effectiveness could be considerably higher. As a result, we investigate how a more effective vaccine would affect the nature of our results. We find that the more effective a vaccine is, the more a central planner would want to deviate from the *ad hoc* allocation (in blue; Figure A19 panels A, B, C, and D). As a result of this greater deviation, we see a larger difference in terms of the reduction in cumulative cases (in red; Figure A19 panels A, B, C, and D). Because a higher effectiveness results in a greater deviation, then, everything else equal, the total workability costs are increased relative to the total vaccine costs (Figure A19 panels E, F, G, and H). The differences are more stark in a world where there is compliance to travel restrictions, as noncompliance blurs the spatial heterogeneity across the jurisdictions leading in general to allocations similar to the ad hoc.

**Figure A19:**
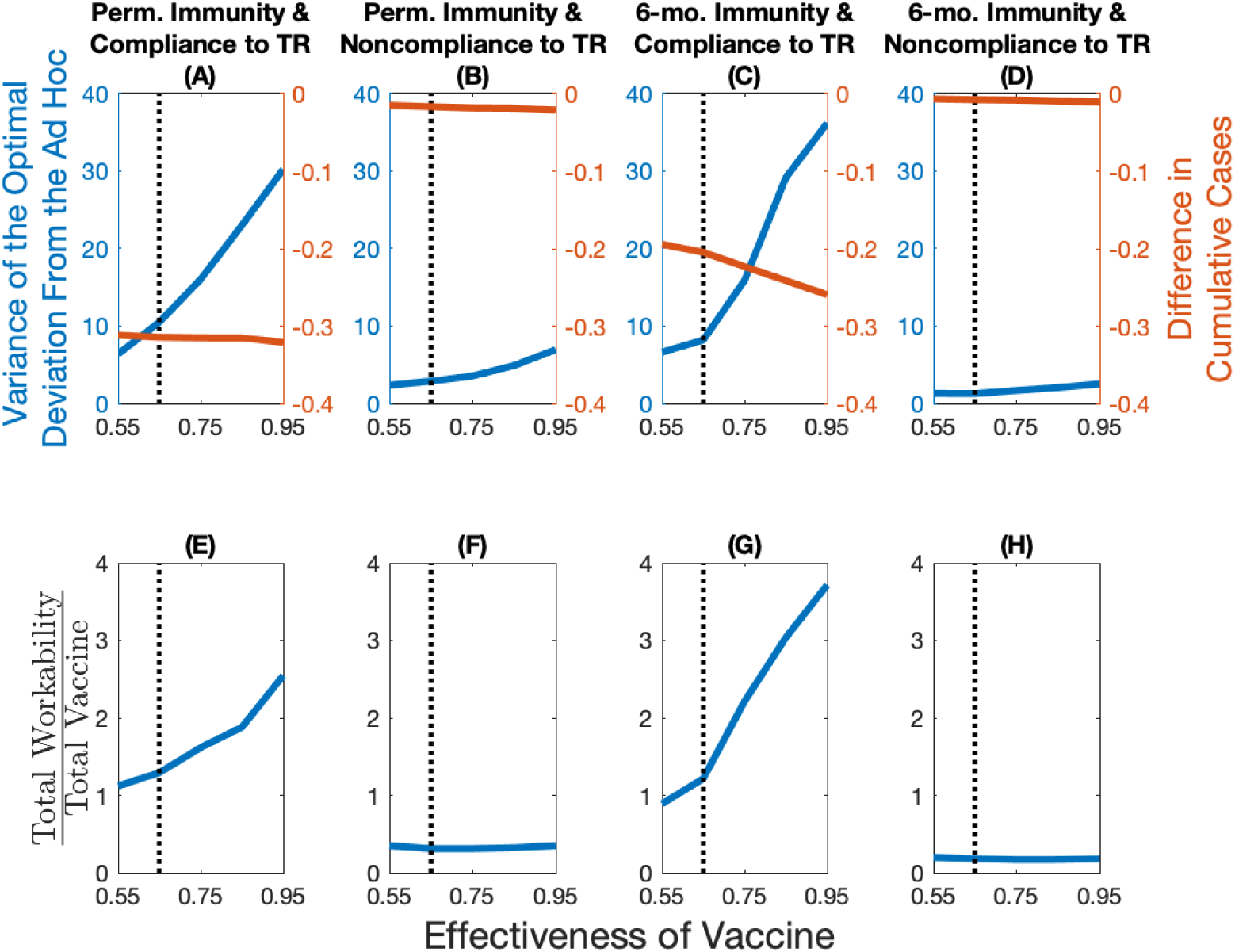
Sensitivity of optimal allocations, and of epidemiological and economic outcomes when varying the effectiveness of the vaccine. The variance of the optimal deviation in percentage (in blue; panels A, B, C, and D) represents an aggregate measure of the optimal deviation from the *ad hoc* allocation. The difference in cumulative cases between the optimal and *ad hoc* allocations (in red; panels A, B, C, and D) represents in percentage terms how well the optimal allocation outperforms the *ad hoc* allocation. The total workability cost over the total vaccine cost (panels E, F, G, and H) represents how many times more the total workability costs are relative to the total vaccine costs. The dotted vertical line in the plots represents the base case value of the vaccine effectiveness (0.65).

## Footnotes

1 See “What value of statistical life does EPA use?” from the U.S. Environmental Protection Agency [9].

2 According to the World Health Organization: “A disability weight is a weight factor that reflects the severity of the disease on a scale from 0 (perfect health) to 1 (equivalent to death).” See: https://www.who.int/healthinfo/global_burden_disease/daly_disability_weight/en/.

3 For more details on how COVID-19’s disability resembles lower respiratory tract infections, see [10].

4 For COVID-19 vaccine prices, see: https://www.npr.org/sections/health-shots/2020/08/06/899869278/prices-for-covid-19-vaccines-are-starting-to-come-into-focus. For a comparison with influenza vaccine prices, see https://www.cdc.gov/vaccines/programs/vfc/awardees/vaccine-management/price-list/index.html.

5 Inspired by the paper of Ryan et al. [11] where the authors show the implications of policy adjustment costs for fisheries management

6 Calibrated using a *R*_0_ estimate from Li et al. [4] and estimates of effects of nonpharmaceutical interventions from Tian et al. [5]; this yields a *R*_0_ of approximately 1.4 when there is compliance to travel restrictions and to match a *R*_0_ of approximately 2.1 when there is no compliance to travel restrictions; these two values representing respectively a “medium” and “low” effect of the nonpharmaceutical intervention.

7 Using estimates from Davies et al. [6]; this represents a 3-day latency period and a 5-day recovery period.

8 Representing a 6-month immunity period in the scenarios where we assume immunity is not permanent; based on [13].

9 Calibrated by using a case-fatility rate of 1.78%. Adjusted for mis- and under-reporting; see [7].

10 Representing the disability associated with severe lower respiratory tract infections because, to our knowledge, there are no official disability estimates associated with COVID-19; see [10].

11 Following Buckner et al. [14], we base this parameter value on the efficiency of the influenza vaccine [15]. Note that the Department of Health and Human Services, Food and Drug Administration (FDA), and Center for Biologics Evaluation and Research (CBER) [16] requires that a future COVID-19 vaccine must have an effectiveness of at least 50%.

12 Based on results from John et al. [12] that suggest a yearly discount rate between 0.3% and 1.5% for health related expenditures; we chose a 1.5% annual discount rate in the main set of results. This gives a monthly discount rate of *r* = 0.0013.

13 Assuming an individual requires two doses; based on current agreements between the U.S. federal government and biotech companies; see https://www.npr.org/sections/health-shots/2020/08/06/899869278/prices-for-covid-19-vaccines-are-starting-to-come-into-focus. For a list of current vaccine prices, and particularly the price of the influenza vaccine, see https://www.cdc.gov/vaccines/programs/vfc/awardees/vaccine-management/price-list/index.html.

14 Value based on a certain proportion of the value of statistical life, *c*; in the base case we assume it is 2 orders of magnitude smaller.

15 Represents a value of statistical life of 10M U.S. dollars. Based on the value of a statistical life that the U.S. Environmental Protection Agency [9] uses: approximately $7.4 million ($2006) which is equivalent to approximately $9.54 million ($2020).

16 The variance is calculated as Var 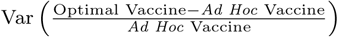. Note that the variance of the optimal deviation from the *ad hoc* is identical in absolute and relative terms across jurisdictions.

